# Longitudinal changes and key determinants of meeting WHO recommended levels of physical activity during the COVID-19 pandemic in a UK-based sample: Findings from the HEBECO Study

**DOI:** 10.1101/2021.09.30.21264358

**Authors:** JJ Mitchell, SJ Dicken, D Kale, A Herbec, E Beard, L Shahab

## Abstract

**Background:** The COVID-19 pandemic has seen government enforced restrictions on movement, with ‘stay-at-home’ orders in place for a second and third time in many nations. Such restrictions have altered physical activity (PA) habits. This study aimed to evaluate longitudinal trends in PA in a self-selected UK-based sample and the key predictors of these trends.

**Methods:** From 23 April 2020 to 30 January 2021, an online survey collected detailed measures of PA engagement in a sample of 1,947 UK-based adults. Generalised estimating equations (GEE) were fitted to explore trends in PA engagement over time, and how sociodemographic, health, lifestyle and contextual factors impacted participant’s attainment of Word Health Organisation (WHO) recommended levels of PA (constituting muscle strengthening activity (MSA), and moderate or vigorous PA (MVPA)).

**Results:** Attainment of WHO recommended levels of total PA showed a decline. While one in five achieved the recommended levels of total PA in the first UK lockdown in April-June 2020 (19.5%, 95%, CI 17.8-21.3%) and a similar proportion in June-July 2020 (17.7%, 95%CI 16.1-19.5%), this reduced significantly during the period of eased restrictions in August-September 2020 (15.2%, 95%CI 13.7-16.9%) and the second UK lockdown in November 2020-January 2021 (14.1%, 95%CI 12.6-15.9%). The same trends were observed for MSA and MVPA individually. Better quality of life, higher socioeconomic position and higher pre-COVID-19 PA levels were associated with meeting WHO recommended PA levels, while having overweight or obesity, a limiting chronic health condition, or being in strict isolation showed the inverse associations. Time-specific associations with MSA or MVPA were observed for gender, age, ethnicity, and other health behaviours.

**Conclusion:** Among a self-selected sample of UK-based adults, there were reductions in PA levels throughout the first UK lockdown without reversal during the ensuing period. Based on observed associations of reduced PA with socioeconomic and health-related indices, such changes may point towards deepening health inequities during the pandemic.

## Background

Most European nations have undergone secondary or tertiary periods of lockdown in early 2021, with state mandated reductions in social contact, outdoor activities, and freedoms in place to reduce the spread of severe acute respiratory syndrome coronavirus 2 (SARS-CoV-2) [1]. One likely consequence of lockdowns has been changes to physical activity (PA) levels. Early evidence from manufacturers of wearable activity-trackers indicates a dose-response effect between the strictness of lockdown measures enacted by regional governments and the severity of reductions in step-count [2, 3]. Since these early findings, an abundance of cross-sectional studies has revealed highly heterogenous changes in PA levels, with some individuals showing improvements in PA levels [4-6] despite an overall downturn in PA [7-16]. This study aimed to corroborate and extend these findings beyond the first strict lockdown and period of eased restrictions in the UK, using an ongoing longitudinal cohort study, the HEalth BEhaviours during the COVID-19 pandemic (HEBECO) study.

Evidence from UK based studies aligns with global trends, showing decreases in individual’s PA across the first lockdown [2, 7, 17-19]. However, early longitudinal studies show mixed evidence of a reversal of this downturn beyond the first strict lockdown [2, 7, 17-19]. Evidence in the UK is not congruous, and the use of varied measures of PA between studies hampers recognition of clinically important changes in PA.

The health consequences of physical inactivity and sedentary behaviours are serious. The World Health Organisation (WHO) recently named physical inactivity as the fourth largest cause of global disease burden and published recommendations for weekly PA, with a view to reducing this burden [20-22]. This encompasses engagement in 150 minutes of MVPA weekly (i.e., activities that increase heart rate and make one feel warmer) and two sessions of MSA per week (e.g., strength/resistance training) [20].

Adherence to the guidelines has been shown to reduce cardiovascular disease risk and mortality associated with physical inactivity [22-28]. Increasing PA engagement can also reduce engagement in harmful behaviours [28-34] while preserving mental and physical wellbeing [29, 35]. Indeed, health behaviour changes are often ‘clustered’, e.g. engaging in less exercise is also associated with increased sedentary time and an increase in health risk behaviours such as alcohol consumption and poorer dietary habits [29]. As such, understanding groups at-risk of poorer PA engagement is a necessary step to unveiling the wider health consequences of enforced lockdowns.

Early evidence has also identified predictors of early negative changes in PA habits. These include older age [12, 13, 36] being female [12, 13], comingfrom a more disadvantaged [12, 36, 37] or non-white background [12, 36], having lower educational level [36], and having poorer physical and mental health [36, 38]. Few studies to date have provided longitudinal evidence beyond the period of eased restrictions, using the framework of WHO recommendations for PA [37, 38] that *i)* corroborates the persistence of associations with these sociodemographic, health and lifestyle factors; *ii)* examines whether poor PA engagement is reversed after lockdown is eased; and *iii)* whether any improvement of PA habits during the inter-lockdown periods is lost during subsequent lockdowns. This study therefore aimed to address this gap, posing the key research questions:

***RQ1***. How has the proportion of individuals attaining WHO recommended levels of total PA, MVPA and MSA, changed since the strict lockdown in April 2020 until January 2021 in a self-selected sample of UK adults?

***RQ2***. Across the period of follow-up, which health, sociodemographic and COVID-19 related situational factors are associated with meeting *i)* WHO recommended levels of total PA, *ii)* WHO recommended levels of MVPA and *iii)* WHO recommended levels of MSA in a self-selected sample of UK adults?

## Methods

### Study Design

The HEBECO study is a longitudinal study of health behaviours during the COVID-19 pandemic in the UK. Recruitment into the baseline wave commenced on 30th April until 14^th^ June 2020. Recruitment was conducted by snowballing via mailing lists at universities and partner organisations. UK-based adults were specifically targeted by advertising on social media platforms and mailing lists for local councils, parliamentary groups related to health, trade unions, sports clubs, charities, and health volunteering groups. The HEBECO cohort has collected data on PA at four different time points, wave 1 in April-May 2020, wave 2 in June-July 2020, wave 3 in August-September 2020 and wave 4 in November 2020-January 2021. Each of these time points has unique situational differences. Wave 1 (April-May 2020; baseline wave) was during the first strict UK lockdown, when non-essential businesses were closed and with a ban on inter-household mixing was in place. Wave 2 (June-July 2020; first follow-up wave) and wave 3 (August-September 2020; second follow-up wave) reflect the return of some individuals to workplaces and some outdoor inter-household mixing in the period of eased restrictions. Wave 4 (November 2020-January 2021; third follow-up wave) was during the second UK lockdown during the early winter and subject to regional differences in restriction severity.

### Participants

The sample included in this study was derived from participants who consented to participate in the HEBECO study. RQ1 only included those participants who contributed the key outcome variables (measures of PA) during the baseline wave (April-June 2020) and wave 3 (August-September 2020), chosen as the two waves of most significance given their starkly different contexts. For RQ2 the sample was further restricted to participants who additionally provided all sociodemographic, health, lifestyle, and COVID-19 situational factors at these same two time points. A final reduced sample of longitudinally complete cases across all waves was used in sensitivity analyses to test for possible bias due to participant attrition.

### Measures

Further details of the measures used are available at https://osf.io/9cmj3/ and within the attached supplementary document.

#### Outcome Variables

Weekly MSA sessions was measured at each time point with the question, ‘*In the past month, on average, on how many days per week have you performed strength training*?’, Answers were dichotomised into ≥2 sessions vs all others.

Weekly MVPA engagement was derived from weekly session frequency and average session duration. Session frequency was measured using the question, ‘*In the past month, on average, how many times per week have you done 15 minutes or more of moderate or vigorous aerobic physical exercise*?’, with answers ranging from ‘0’ to ‘14 or more’ in single frequency increments. This upper limit constraint, set to 2 sessions per day on average is in line with previous output from this cohort [11]. Average session duration was measured with the question, ‘*In the past month, how long (in minutes) was your average session of moderate or vigorous aerobic physical activity?*’. Answers were recorded on an interval scale ranging from ‘15 minutes’ to ‘480 minutes or more’ in 15 minute increments. The upper limit constraint of 8 hours capped highly atypical responses.

Meeting WHO recommended levels of total PA was a binary composite score of meeting recommended levels of both MVPA and MSA vs all others. Pre-COVID-19 exercise levels were also measured retrospectively at baseline in the same manner. All outcome measures were time-varying. A final count measure of PA sessions was utilised in sensitivity analysis and derived by summing the weekly number of MVPA and MSA sessions reported by participants.

#### Explanatory Factors

Explanatory factors were included based on previous evidence during the first COVID-19 lockdown periods in the UK and Europe [12, 13, 36, 37, 39] and encompassed sociodemographic and lifestyle and health factors. Lastly, exploratory COVID-19 situational factors were also included.

#### Sociodemographic factors

Sociodemographic explanatory variables included age (continuous), gender (female vs all others) and ethnicity (white vs all others), measured at baseline and included as fixed (time-invariant) covariates. Socioeconomic position was also time-invariant, measured at baseline using a novel, composite index based on participants educational level, household income, and housing tenure. A sum score ranging from 0-3 was derived where one point was issued for each of i) attainment of at least A-level or equivalent qualifications, ii) house ownership (outright or mortgage), iii) possessing a total household income of ≥£50,000. An employment status/working from home composite measure was time varying. Those in full or part time employment or education were subdivided into those who can work entirely at home and those who must attend their place of work/study.

#### Lifestyle and Health factors

Lifestyle and health explanatory variables included Quality of Life which was time-varying and scores participant living situation, social relationships, family relationships and psychological wellbeing. Perceived Health Risk from COVID-19 was time varying, operationalised as a dichotomy of ‘major to moderate risk’ and ‘minimal to no risk’. Having a Body mass index (BMI; body mass (*kg*) /height (*m*)^2^) of overweight or obese was derived at each time point from baseline height and self-reported body weight at a given wave, dichotomised into overweight or obese (≥25.00) and all others including ‘don’t know’ and ‘prefer not to say’. A weighted mean of body weight at waves 1 and 3 was used to impute BMI at wave 2 where weight was not collected. Having a confirmed or suspected COVID-19 infection was time-varying. Presence of a limiting health conditions which might impact on the participant’s ability to engage in PA was measured at baseline and fixed throughout. Lastly, risky health behaviours including high alcohol consumption (>14 units/week) and current smoking were included as time-varying factors.

#### COVID-19 situational factors

COVID-19 situational factors included isolating and home-environmental factors which may confound on exercise levels throughout this period. Isolation was measured at all time-points and dichotomised into strict isolation vs all others. Adequate exercise space access was fixed, measured at baseline. Lastly, participants were dichotomised into those living alone or with others at each time point.

### Statistical Analysis

Chi-square and *t*-tests were used to compare the participants who fulfilled eligibility criteria (analytic sample) with those in the excluded sample. Analyses were conducted using SPSS 22.0 (IBM, Armonk, NY, USA). For RQ1, time-trend analyses were conducted by fitting generalized estimating equations (GEE) [40] for each outcome (total PA (MVPA and MSA combined) MVPA and MSA respectively) across waves (timepoints), which estimated the proportions of the sample attaining WHO recommended levels of PA without adjustment for covariates. Pairwise comparisons between time points were made using a type III Wald Chi-square test with Šidák correction for family-wise error [40, 41]. Given the binary nature of the outcomes and the repeated measurement of participants, GEE were fitted using the log-link function and using an autoregressive correlation matrix (selected based on study design and best fit, indicated by the Quasilikelihood under the Independence model Criterion (QIC)) [40]. For RQ2, univariate models were first fitted between each factor and outcome variable and included a timepoint*predictor interaction term to explore time-specific effects. A forward selection approach was then used where all predictors and any significant interaction terms which improved final model fit were individually included in the final full model. For nonsignificant predictors, Bayes Factors (BF) were calculated based on previous evidence exploring the impact of sociodemographic characteristics on PA levels, to differentiate no-effect from small effects [42-45].

Missing data were treated as missing at random. The study protocol was pre-registered on the Open Science Framework (OSF) before analysis (https://osf.io/q2zak/). Deviations from the pre-registered protocol are described in the supplement and are available in the supplement.

Sensitivity analyses using longitudinally complete cases were used to check the robustness of findings. Furthermore, a count variable of PA sessions was used to explore whether associations observed in RQ2 were present with an outcome more indicative of total PA levels to aid in comparison with previous literature.

## Results

### Sample Characteristics

The analytic sample of 1,947 UK-based adults was principally female (70%, N=1363), middle aged (mean=50, standard deviation (SD)=14.7) and of white ethnicity (95%, N=1850) (table 1). Our sample also contained a high proportion (30%, N=584) of participants in the highest socioeconomic position based on household income, education, and home ownership relative to the proportions that might be expected from ONS census data [46]. Some significant differences existed between the analytic and excluded samples. The included sample were significantly younger (*p<0*.*001)*, had more females (*p=0*.*024)*, fewer smokers (*p<0*.*001)* and more individuals of overweight or obese BMI *(p<0*.*008)*. Further comparisons are presented in table 1.

**Table 1.** Participant demographics at baseline.

|  | **Analytic**<br>**Sample**<br>**N=1,947** | **Excluded**<br>**Sample**<br>**N=1,045** | **Comparison**<br>**P-value** |
| --- | --- | --- | --- |
| **Mean Age (SD)** | **50.61**<br>**(14.66)** | **42.93**<br>**(15.68)** | **<0.001 \*** |
| **Female Gender, % (N)** | **70% (1363)** | **66% (690)** | **0.024\*** |
| **White Ethnicity, % (N)** | **95% (1850)** | **91% (951)** | **<0.001\*** |
| **Country of residence, % (N)** |  |  | 0.36 |
| England | **86% (1674)** | **85% (888)** |  |
| Scotland | **8% (156)** | **7% (73)** |  |
| Wales | **5% (97)** | **6% (63)** |  |
| Northern Ireland | **1% (19)** | **2% (21)** |  |
| **Composite Socioeconomic Index, % (N)** |  |  | **<0.001\*** |
| Highest | **30% (584)** | **26% (272)** |  |
| Upper | **31% (604)** | **34% (355)** |  |
| Lower | **26% (506)** | **32% (334)** |  |
| Lowest | **3% (59)** | **8% (84)** |  |
| **Employment Status, % (N)** |  |  | **<0.001\*** |
| Employed & Working from Home | **51% (993)** | **52% (543)** |  |
| Employed & Attending Workplace | **18% (350)** | **23% (240)** |  |
| Unemployed, Retired or full-time Carer | **31% (604)** | **25% (261)** |  |
| **Balcony/Garden Access at home % (N)** | **73% (1421)** | **68% (711)** | **0.007\*** |
| **Living Alone % (N)** | **17% (331)** | **16% (167)** | 0.36 |
| **Time-varying characteristics at baseline.** |  |  |  |
| **Smoker % (N)** | **14% (273)** | **28% (293)** | **<0.001\*** |
| **Alcohol Consumption % (N)** |  |  | 0.24 |
| Greater than 14 weekly units | **19% (370)** | **20% (209)** |  |
| Less than or equal to 14 weekly units | **81% (1577)** | **80% (836)** |  |
| **Confirmed or Suspected COVID-19 Infection % (N)** | **20% (389)** | **23% (240)** | **0.041\*** |
| **Strict Isolation % (N)** | **7% (136)** | **7% (73)** | 0.95 |
| **Composite Quality of Life Index (0-5) Mean (SD)** | **3.43 (0.47)** | **3.24 (0.88)** | **<0.001\*** |
| **BMI (Overweight) % (N)** | **54% (1051)** | **50% (523)** | **0.008\*** |
| **Chronic condition or Disability % (N)** | **11% (214)** | **11% (115)** | 0.70 |
| **Meeting WHO Recommended Total PA % (N)** | **20% (389)** | **20% (209)** | 0.86 |
| **Meeting WHO Recommended MVPA % (N)** | **44% (857)** | **41% (428)** | 0.25 |
| **Meeting WHO Recommended MSA % (N)** | **34% (662)** | **35% (366)** | 0.84 |

\*Significant differences in included and excluded sample at $\alpha \leq 0.05$ as indicated by *t*-test of means or Pearson Chi-Square test of difference of proportions ( $\chi^2$ ).

#### RQ1: Meeting WHO Recommended levels of total PA, MVPA and MSA

Time-trend analysis demonstrated a steady decline in the proportions of individuals meeting WHO recommended levels of all PA types across the study period (figure 1). The proportion of individuals attaining WHO recommended levels of total PA decreased from baseline wave 1 (April-May) (19.5%, 95% CI 17.8-21.3%) to wave 2 (June-July) (17.7%, 95% CI 16.1-19.5%), wave 3 during the period of eased restriction (Aug-Sept) (15.2%, 95% CI 13.7-16.9%) and wave 4 during the second UK lockdown (Nov-Jan) (14.1%, 95% CI 12.6-15.9%). However, no significant differences were observed between the first two or latter two waves. Both MVPA and MSA showed similar decreases across the study period, but again showed no significant differences between waves 1 and 2, or between waves 3 and 4 (figure 1). This trend persisted in complete case analysis (supplementary tables 6,7 and 8).

**Figure 1.**
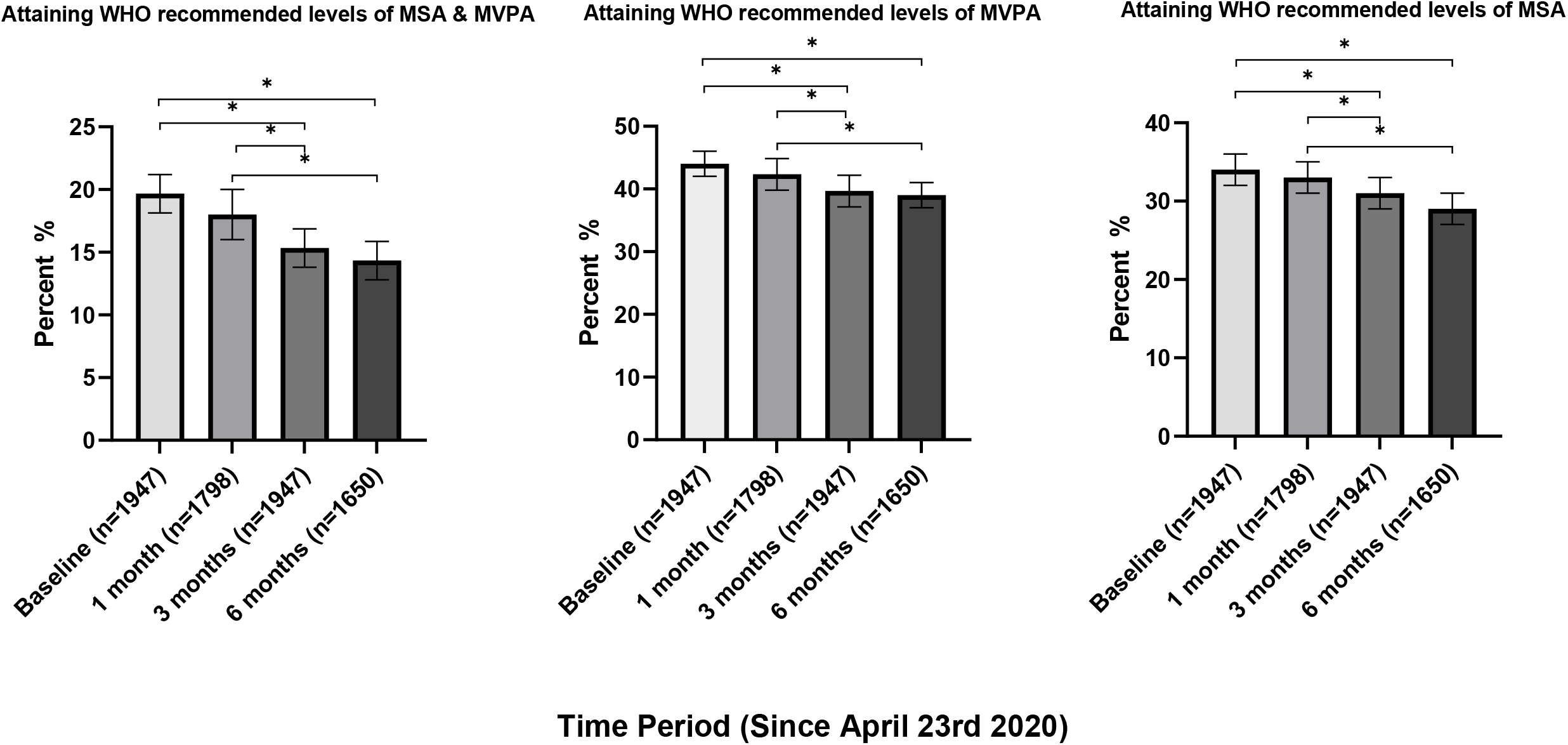
Time trend analysis of PA levels across the COVID-19 pandemic. Time-trends in proportions of participants meeting MVPA, MSA and both WHO PA guidelines & Wald χ2 pairwise comparison with Šidák correction. *p < 0.05

#### RQ2 i) Predictors of meeting WHO recommended levels of total PA

Sociodemographic and situational factors associated with meeting WHO recommended levels of total PA included socioeconomic position, being in strict isolation and those living alone. The highest two socioeconomic positions were associated with attaining WHO recommended levels of total PA (table 2). Those in strict isolation showed lower odds of meeting WHO recommended levels of total PA. Living alone showed time-specific effects only, predicting higher total PA attainment specifically during the period of eased restriction in August-September (*p=0*.*001)* (Figure 2; supplementary table 1). Health and behavioural factors including having overweight or obesity and a higher perceived risk from COVID-19 infection were negatively associated with attaining total recommended levels of total PA (table 2). The biggest predictors of meeting WHO recommended levels of total PA during the study period was meeting them prior to the COVID-19 pandemic (table 2). This relationship with pre-COVID PA levels also showed significant moderation by time, showing stronger associations by the second lockdown (*p=0*.*037*) and suggesting that failing to meet recommended PA levels pre-pandemic became more strongly associated with not meeting total PA levels as the pandemic progressed (Figure 2; supplementary table 2). Inclusion of time itself in adjusted models confirmed the unadjusted time-trend analysis, as time showed incrementally lower odds of meeting all PA types further into the pandemic (supplementary tables 1-5).

**Table 2.**
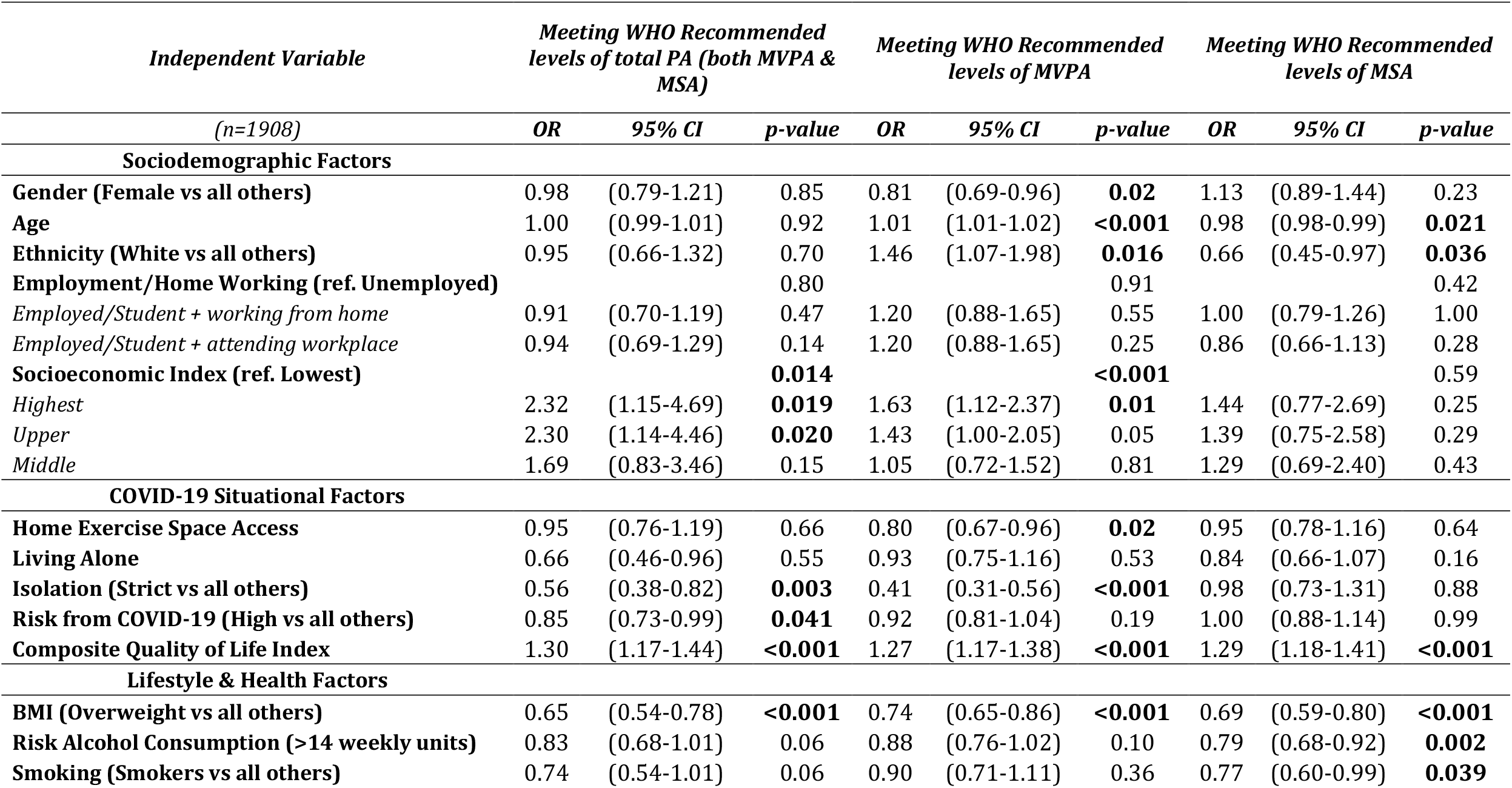
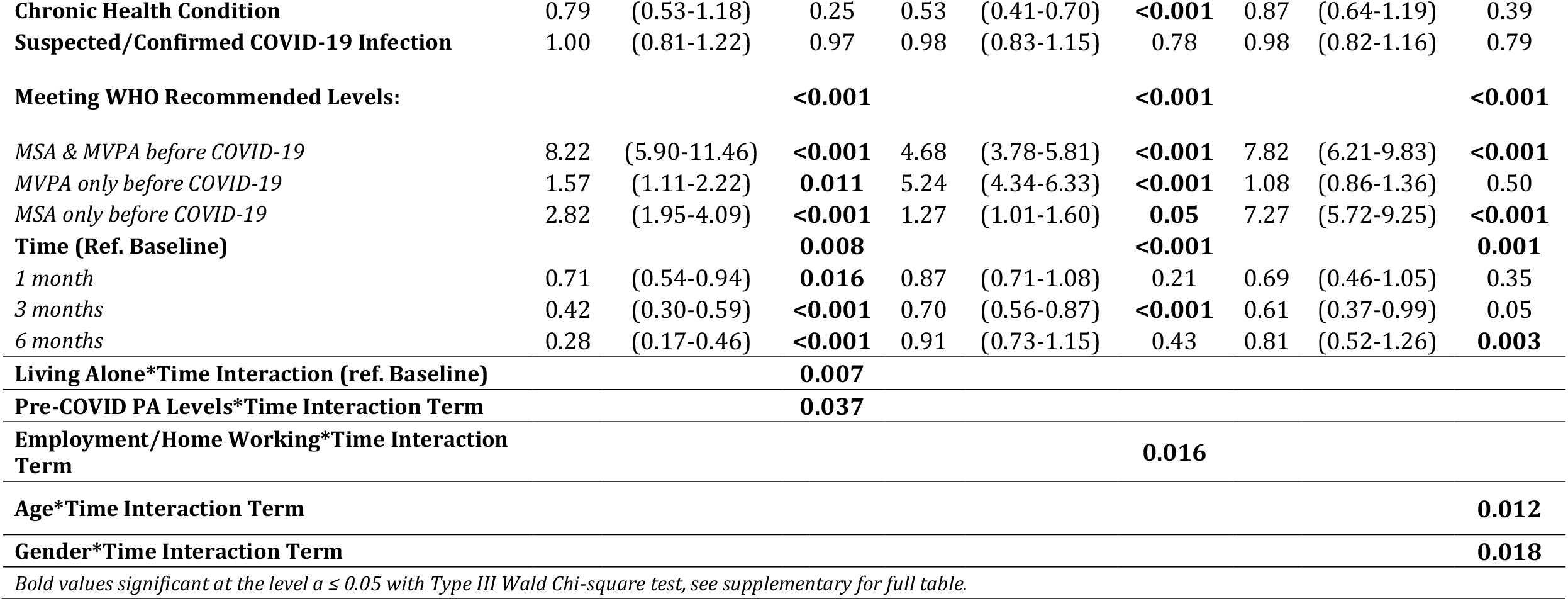
Factors associated with meeting WHO recommended levels of PA throughout the COVID-19 Pandemic.

| *Independent Variable*<br>(*n=1908*) | *Meeting WHO Recommended*<br>*levels of total PA (both MVPA &*<br>*MSA)* |  |  | *Meeting WHO Recommended*<br>*levels of MVPA* |  |  | *Meeting WHO Recommended*<br>*levels of MSA* |  |  |
| --- | --- | --- | --- | --- | --- | --- | --- | --- | --- |
|  | *OR* | *95% CI* | *p-value* | *OR* | *95% CI* | *p-value* | *OR* | *95% CI* | *p-value* |
| **Sociodemographic Factors** |  |  |  |  |  |  |  |  |  |
| **Gender (Female vs all others)** | 0.98 | (0.79-1.21) | 0.85 | 0.81 | (0.69-0.96) | **0.02** | 1.13 | (0.89-1.44) | 0.23 |
| **Age** | 1.00 | (0.99-1.01) | 0.92 | 1.01 | (1.01-1.02) | **<0.001** | 0.98 | (0.98-0.99) | **0.021** |
| **Ethnicity (White vs all others)** | 0.95 | (0.66-1.32) | 0.70 | 1.46 | (1.07-1.98) | **0.016** | 0.66 | (0.45-0.97) | **0.036** |
| **Employment/Home Working (ref. Unemployed)** |  |  | 0.80 |  |  | 0.91 |  |  | 0.42 |
| *Employed/Student + working from home* | 0.91 | (0.70-1.19) | 0.47 | 1.20 | (0.88-1.65) | 0.55 | 1.00 | (0.79-1.26) | 1.00 |
| *Employed/Student + attending workplace* | 0.94 | (0.69-1.29) | 0.14 | 1.20 | (0.88-1.65) | 0.25 | 0.86 | (0.66-1.13) | 0.28 |
| **Socioeconomic Index (ref. Lowest)** |  |  | **0.014** |  |  | **<0.001** |  |  | 0.59 |
| *Highest* | 2.32 | (1.15-4.69) | **0.019** | 1.63 | (1.12-2.37) | **0.01** | 1.44 | (0.77-2.69) | 0.25 |
| *Upper* | 2.30 | (1.14-4.46) | **0.020** | 1.43 | (1.00-2.05) | 0.05 | 1.39 | (0.75-2.58) | 0.29 |
| *Middle* | 1.69 | (0.83-3.46) | 0.15 | 1.05 | (0.72-1.52) | 0.81 | 1.29 | (0.69-2.40) | 0.43 |
| **COVID-19 Situational Factors** |  |  |  |  |  |  |  |  |  |
| **Home Exercise Space Access** | 0.95 | (0.76-1.19) | 0.66 | 0.80 | (0.67-0.96) | **0.02** | 0.95 | (0.78-1.16) | 0.64 |
| **Living Alone** | 0.66 | (0.46-0.96) | 0.55 | 0.93 | (0.75-1.16) | 0.53 | 0.84 | (0.66-1.07) | 0.16 |
| **Isolation (Strict vs all others)** | 0.56 | (0.38-0.82) | **0.003** | 0.41 | (0.31-0.56) | **<0.001** | 0.98 | (0.73-1.31) | 0.88 |
| **Risk from COVID-19 (High vs all others)** | 0.85 | (0.73-0.99) | **0.041** | 0.92 | (0.81-1.04) | 0.19 | 1.00 | (0.88-1.14) | 0.99 |
| **Composite Quality of Life Index** | 1.30 | (1.17-1.44) | **<0.001** | 1.27 | (1.17-1.38) | **<0.001** | 1.29 | (1.18-1.41) | **<0.001** |
| **Lifestyle & Health Factors** |  |  |  |  |  |  |  |  |  |
| **BMI (Overweight vs all others)** | 0.65 | (0.54-0.78) | **<0.001** | 0.74 | (0.65-0.86) | **<0.001** | 0.69 | (0.59-0.80) | **<0.001** |
| **Risk Alcohol Consumption (>14 weekly units)** | 0.83 | (0.68-1.01) | 0.06 | 0.88 | (0.76-1.02) | 0.10 | 0.79 | (0.68-0.92) | **0.002** |
| **Smoking (Smokers vs all others)** | 0.74 | (0.54-1.01) | 0.06 | 0.90 | (0.71-1.11) | 0.36 | 0.77 | (0.60-0.99) | **0.039** |
| **Chronic Health Condition** | 0.79 | (0.53-1.18) | 0.25 | 0.53 | (0.41-0.70) | <**0.001** | 0.87 | (0.64-1.19) | 0.39 |
| **Suspected/Confirmed COVID-19 Infection** | 1.00 | (0.81-1.22) | 0.97 | 0.98 | (0.83-1.15) | 0.78 | 0.98 | (0.82-1.16) | 0.79 |
| **Meeting WHO Recommended Levels:** |  |  | <**0.001** |  |  | <**0.001** |  |  | <**0.001** |
| *MSA & MVPA before COVID-19* | 8.22 | (5.90-11.46) | <**0.001** | 4.68 | (3.78-5.81) | <**0.001** | 7.82 | (6.21-9.83) | <**0.001** |
| *MVPA only before COVID-19* | 1.57 | (1.11-2.22) | **0.011** | 5.24 | (4.34-6.33) | <**0.001** | 1.08 | (0.86-1.36) | 0.50 |
| *MSA only before COVID-19* | 2.82 | (1.95-4.09) | <**0.001** | 1.27 | (1.01-1.60) | **0.05** | 7.27 | (5.72-9.25) | <**0.001** |
| **Time (Ref. Baseline)** |  |  | **0.008** |  |  | <**0.001** |  |  | **0.001** |
| *1 month* | 0.71 | (0.54-0.94) | **0.016** | 0.87 | (0.71-1.08) | 0.21 | 0.69 | (0.46-1.05) | 0.35 |
| *3 months* | 0.42 | (0.30-0.59) | <**0.001** | 0.70 | (0.56-0.87) | <**0.001** | 0.61 | (0.37-0.99) | 0.05 |
| *6 months* | 0.28 | (0.17-0.46) | <**0.001** | 0.91 | (0.73-1.15) | 0.43 | 0.81 | (0.52-1.26) | **0.003** |
| **Living Alone\*Time Interaction (ref. Baseline)** |  |  | **0.007** |  |  |  |  |  |  |
| **Pre-COVID PA Levels\*Time Interaction Term** |  |  | **0.037** |  |  |  |  |  |  |
| **Employment/Home Working\*Time Interaction Term** |  |  |  |  |  | **0.016** |  |  |  |
| **Age\*Time Interaction Term** |  |  |  |  |  |  |  |  | **0.012** |
| **Gender\*Time Interaction Term** |  |  |  |  |  |  |  |  | **0.018** |
| *Bold values significant at the level $\alpha \leq 0.05$ with Type III Wald Chi-square test, see supplementary for full table.* |  |  |  |  |  |  |  |  |  |

**Figure 2.**
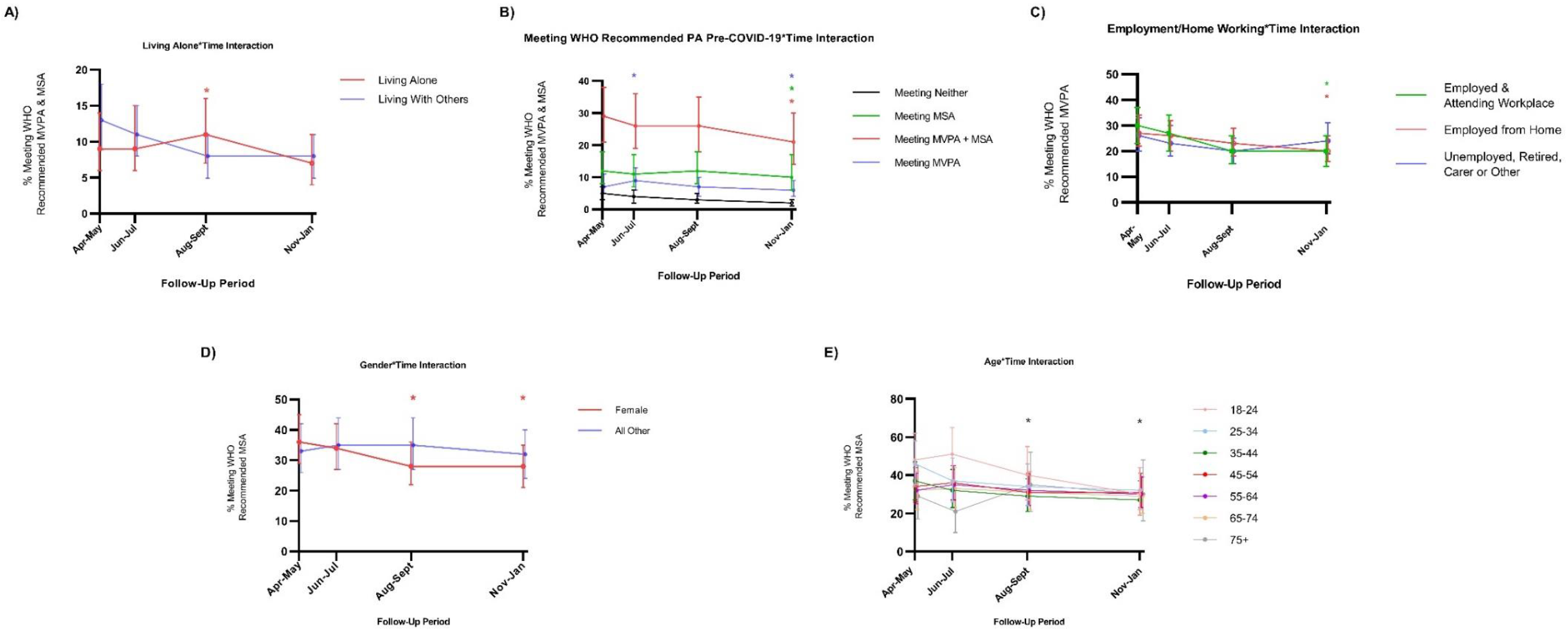
Time-varying associations of sociodemographic factors & meeting WHO recommended levels of PA. Time-differential associations of factors with meeting total WHO recommended PA levels, MSA & MVPA. A) Meeting any PA recommendation pre-COVID-19 increased odds of meeting WHO recommended levels of PA during the second lockdown. B) Living alone showed higher odds of meeting total WHO recommended levels of PA during the period of eased restriction. C) Working/studying from home or attending a workplace was associated with higher odds of attainment of MVPA during the second lockdown. D) Gender (female) showed lower odds of MSA attainment during the period of eased restriction and second lockdown. E) Age shows a positive association with MSA attainment at these time points. * p<0.05

#### RQ2 ii) Predictors of meeting WHO recommended levels of MVPA

Sociodemographic and situational factors including age, gender, white ethnicity, and employment status showed varying associations with meeting WHO recommended levels of MVPA. Employment status showed time-specific associations with attaining sufficient MVPA. Both being employed from home (*p=0*.*039*) or attending a workplace (*p=0*.*020*) was associated with less MVPA engagement compared with being unemployed, retired, or a full-time carer during the second UK lockdown in November-January (Figure 2; supplementary table 3). Gender (female) showed a substantial negative association with MVPA attainment while white ethnicity showed a positive association (table 2). Unusually, having access to adequate exercise space at home was negatively associated with attaining MVPA levels. Again, health and behavioural factors showed significant negative associations with MVPA attainment. Participants with an overweight or obese BMI showed lower odds of attaining recommended levels of MVPA as did those with a chronic health condition. Pre-COVID-19 PA engagement and higher quality of life again proved the strongest predictors of attaining sufficient MVPA throughout the study period (table 2).

#### RQ2 iii) Predictors of meeting WHO recommended levels of MSA

Sociodemographic factors including age and being female both showed time-differential, negative effects on MSA at both wave 3 in August-September (*p=0*.*008; p=0*.*002*) and wave 4 in November-January (*p=0*.*001; p=0*.*027*) (Figure 2; supplementary table 5). Participants of white ethnicity showed substantially reduced odds of attaining recommended levels of MSA, as too did participants with an overweight or obese BMI (table 2). Risky health behaviours showed associations specifically with MSA engagement. Participants consuming alcohol above the recommended weekly limits as well as smokers both showed lower odds of meeting recommended levels of MSA (table 2). Lastly, higher quality of life and attaining sufficient MSA pre-COVID-19 proved strong predictors of meeting recommended MSA levels during the study period.

### Nonsignificant Findings

Despite associations between gender, age and having a chronic health condition and the individual aspects of the PA recommendations, there was no significant findings for an association between these factors and meeting total WHO recommended levels of PA. Bayes Factors were calculated based on previous evidence reporting a negative effect of a chronic health condition (OR: 0.56–0.96) and positive effect of gender (female) (OR=1.10 to 2.33). BF’s showed there was insufficient evidence to rule out an association between having a chronic health condition and meeting WHO recommended levels of PA (BF=0.88). Data were also insensitive for gender (BF=1.06). For age however, our data provided evidence of no effect (BF=0.01) [16, 36].

### Sensitivity Analysis

To assess for possible bias caused by attrition, final models were completed with complete cases only (supplementary table 14-16). Trends in PA were robust (supplementary tables 9-11), as were the significance and directions of associations in all models except MSA models, in which alcohol consumption and ethnicity lost significance. To aid comparability of this study with previous literature, and account for the limited sensitivity of the WHO recommendations for total PA engagement, analyses were repeated using a count of weekly PA sessions. Time-trends showed substantive differences at the level of PA sessions (supplementary table 17). Perceived risk from COVID-19, alcohol consumption and smoker status no longer showed significant associations in any model, nor did the time-specific effects of living alone and working from home; however. all other findings remained robust at the level of total PA measured on a linear scale, despite changes to point estimates (supplementary tables 18-22).

## Discussion

Here we provide evidence for clinically meaningful changes in the PA levels of a self-selected UK-based sample of adults throughout four unique contexts of the COVID-19 pandemic. Proportions of individual’s meeting WHO recommended levels of total PA, MVPA and MSA all showed a decrease throughout April 2020 to January 2021; however, unlike previous studies which examined total PA habits, no recovery was observed during the period of eased restrictions in August-September 2020 [37, 38]. This study identified no significant difference between wave 1 during the first strict lockdown in April-May and wave 2 at the ending of the first lockdown in June-July. However, this study extends previous literature, identifying significant drops between these waves and the August-September wave during the period of eased restriction and November-January wave during the second lockdown in the UK [2, 47, 48]. In general, MVPA attainment in this sample was far lower than might be expected from other estimates of PA in the UK population [49, 50].

Many sociodemographic factors were predictive of differential PA engagement. White ethnicity was associated with meeting recommended levels of MVPA, but negatively associated with MSA attainment. Previous studies have observed higher attainment of both MVPA and MSA in white communities [51]. This finding suggests that individuals from different backgrounds may be opting to exercise in different ways during the lockdown.

Despite Bayes Factors suggesting inconclusive evidence for an effect of gender on meeting the WHO guideline levels of total PA, our data does still raise possible sex differences and socioeconomic inequities in attaining sufficient levels of MVPA and MSA. Females showed lower odds of meeting recommended levels of MVPA, specifically during the period of eased restriction and second lockdown. Increasing age showed the inverse associations with MVPA and MSA in line with other studies, but Bayes Factors provided evidence for ‘no effect’ of age on meeting WHO recommended PA levels [36]. Evidence is mixed as to the existence, and possible mediators, of sex-differences in PA engagement during the pandemic [16, 52]. Indeed, early cross-sectional data in other UK cohorts indicated significantly higher attainment of MVPA in females [16]; however, subsequent longitudinal studies have suggested little or no difference [53]. Age-related differences have been observed almost unanimously [16, 37] and may be driven by factors such as additional time and freedoms afforded to those who are financially independent which often accompanies older age [54]. However, our study does not corroborate this finding associated with age, despite still identifying strong relationships between greater PA engagement and socioeconomic position and high quality of life [55]. Further to this, participants engaged in negative health behaviours including smoking and excessive alcohol consumption were at greater risk of insufficient PA. These trends therefore align with previous evidence, which suggests the existence of ‘clustering’ of negative health behaviours and an exacerbation of sex-differences and socioeconomic inequities in PA engagement during the COVID-19 pandemic. [29].

The most at-risk groups for poor PA engagement included those with a chronic health condition (e.g. stroke, heart disease, dementia), with overweight or obesity and those with a higher perceived risk from COVID-19, suggesting greater efforts should be made to enable those individuals asked to shield to complete their regular PA, especially given the consequences of even transiently increased sedentarism may be more severe for those with chronic conditions [22, 24, 56-58].

Situational factors were also associated with differing PA engagement. Living alone proved a positive predictor of meeting WHO recommended levels of total PA, specifically during the period of eased restrictions in August-September 2020, perhaps highlighting the importance of social sports and gyms for these individuals to attain their PA. Having adequate space indoors in which to exercise was negatively associated with attaining adequate MVPA. Perhaps expectedly therefore, MVPA proved the component of participant’s PA requirements most attenuated by being confined to the indoors whilst having to strictly isolate. This effect size was reduced when looking at total PA sessions, implying that indoor MVPA sessions may be fewer, shorter, or more difficult to attain than MSA, opening the possibility that MSA interventions may prove a useful and more sustainable target for individuals required to shield.

Lastly, pre-COVID-19 PA levels were the most consistent predictors of attainment of recommended levels of all PA types, suggesting relatively strong habits regarding routine PA. Establishing sufficient PA engagement more widely in the wake of the COVID-19 pandemic may therefore remain the optimal target for primary prevention of poor PA attainment during future lockdowns.

## Strengths and Limitations

This study provides novel evidence on factors associated with differences in PA engagement across four unique contexts during the pandemic in the UK and holistically explored the factors which may impact PA engagement, using a framework of internationally accepted recommendations for clinically meaningful PA.

Significant limitations, however, do exist. The WHO guidelines for PA are binary, thus not reflecting the total daily energy expenditure which is known to be a significant modulator of the beneficial effects of PA [22]. Despite repeating analyses with a count variable of PA session frequency, this study is not immediately comparable with studies examining total PA. Sedentary time, which has been shown to have independent, negative health effects [22], was not measured. Further studies should address this with objectively measured PA and sedentary time, while adopting the WHO recommendations as a standard.

The period of eased restriction and subsequent lockdowns beyond the first strict UK-wide lockdown varied by region, and thus individuals may have had differing restrictions during wave 4 depending on their region, as well as differing seasonal and weather trends, which were not modelled in this analysis. Several factors were necessarily coded as binary, including alcohol consumption and smoking, which does not account for the significant heterogeneity within these health behaviours. Lastly, use of a self-selected sample, the demographics of which do not accurately represent those of the wider UK population, and the reliance on self-reported measures opens the opportunity for both sampling and social desirability biases.

## Conclusion

This study used PA data contributed by a self-selected UK-based sample to measure population level estimates and possible barriers to meeting WHO recommended levels of PA engagement over an eight-month period since April 2020. We observed population-level decreases in PA across the study period, without recovery during the periods outside of lockdown. Several at-risk groups were also identified with consistently lower odds of meeting the WHO recommended levels of PA, which could lead to greater health inequities over time. Future research should make efforts to further discern the situational and motivational reasons behind differences in these at-risk groups to inform future interventions and PA promotion.

## Data Availability

Data are available upon request.

## Abbreviations

COVID-19: Coronavirus SARS-COV2 disease 2019
SARS-CoV-2: Severe acute respiratory syndrome coronavirus 2
SARS: Severe acute respiratory syndrome
PA: Physical Activity
MVPA: Moderate or Vigorous Physical Activity
MSA: Muscle Strengthening Activity
WHO: World Health Organisation

## Acknowledgements

The team would like to pay sincere thanks to the participants in this study for their contribution during the ongoing pandemic.

## Author Contributions

AH and LS conceived the study and designed the survey. JJM, SD and EB conducted analysis and interpretation. JJM drafted the manuscript. All authors examined final analyses and revised several drafts before approving the final manuscript.

## Appendix A. Supplementary data

Supplementary data for this article is included with this submission.

Study protocol and survey documents can be found online at: Home | UCL HEBECO STUDY(ucl-covid19research.co.uk).

## Ethical Approval

The study has been approved by UCL Research Ethics Committee at the UCL Division of Psychology and Language Sciences (PaLS) (CEHP/2020/579) as part of the larger programme ‘The optimisation and implementation of interventions to change behaviours related to health and the environment’.

