## Supplementary File for "Longitudinal changes and key determinants of meeting WHO recommended levels of physical activity during the COVID-19 pandemic in a UK-based sample: Findings from the HEBECO Study"

**Supplementary Materials**

#### 1. Substantive deviations from the pre-registered protocol

The pre-registered protocol may be obtained at OSF: https://osf.io/q2zak/

i) Choice of covariance matrix was not decided based on the fit of Akaike's Information Criterion, but Quasi-likelihood under the Independence model Criterion (QIC), decided based on best practise with performing Generalised Estimating Equations (GEE) analysis. This was assessed on each outcome with only time as an independent variable (IV). QIC was reassessed on final models with different correlation matrices and including all predictors. This confirmed the choice of autoregressive correlation matrix as the most suitable.

ii) Model building no longer followed sequential adjustment initially set-out. Rather, for RQ2 we first modelled each IV, only including time. We next trialled a time*IV interaction. Lastly we included all IV's and any significant IV*time interactions in the final model and removed interactions based on substantive improvements to QIC. This was repeated for each of the outcomes.

iii) Smoking and alcohol consumption was not originally included in the pre-registered protocol. Inclusion of smoking was reconsidered based on evidence of substantial changes to smoking throughout the same period of the pandemic using the HEBECO cohort (Kale et al., 2021). Given the clustering of health behaviours, changes to smoking, and to alcohol consumption throughout the period were therefore considered valuable predictors of trends in other health behaviours, the probable meaningful cardiovascular implications of which were considered pertinent and hence included.

iv) Lastly weighting of data was only used for descriptive purposes and was not reported. Weighting of data was later not included in any inferential statistics for the sake of the accuracy of results. Previous outputs from the HEBECO study's large sample have reported data unweighted in the knowledge that the sample is self-selected and reported as such. Hence unweighted data were used in analyses, improving accuracy of results at the cost of generalizability.

#### 2. Full tables from all reported analyses are presented below.

#### 3. Detailed descriptions of how the measures were collected and recoded is outlined at the end of this supplementary document.

### Supplementary Tables Contents

**Tables Contents** ........ 2

**Meeting MVPA & MSA Full tables**

Supplementary table 1. Meeting MVPA & MSA levels (Living alone*Time) ........ 4

Supplementary table 2. Meeting MVPA & MSA levels (Pre-COVID-19 PA*Time) ........ 6

**Meeting MVPA Full tables**

Supplementary table 3. Meeting MVPA levels (Employment/Home Working*Time) ........ 8

**Meeting MSA Full tables**

Supplementary table 4. Meeting MSA levels (Gender*Time) ........ 10

Supplementary table 5. Meeting MSA levels (Age*Time) ........ 12

**Pairwise Comparisons (Total Sample**)

Supplementary table 6. Meeting MVPA & MSA Pairwise comparisons ........ 15

Supplementary table 7. Meeting MVPA Pairwise comparisons ........ 16

Supplementary table 8. Meeting MSA Pairwise comparisons ........ 17

**Complete Case Analysis**

Supplementary table 9. Meeting MVPA & MSA Pairwise comparisons (Complete Case) ........ 18

Supplementary table 10. Meeting MVPA Pairwise comparisons (Complete Case) ........ 19

Supplementary table 11. Meeting MSA Pairwise comparisons (Complete Case) ........ 20

Supplementary table 12. Meeting MSA levels (Gender*Time & Age*Time Combined) ... 21
Supplementary table 13. Meeting MVPA & MSA levels (PA*Time & Living alone*Time Combined) ... 23

**Complete Case Analysis**

Supplementary table 14. Meeting MVPA & MSA Full Models (Complete Case) ... 26
Supplementary table 15. Meeting MVPA Full Models (Complete Case) ... 28
Supplementary table 16. Meeting MSA Full Models (Complete Case) ... 30

**Complete Case Analysis**

Supplementary table 17. PA Session Count Variable Pairwise comparisons ... 31
Supplementary table 18. PA Session Count Variable 'Meeting WHO Total PA Pre-COVID'*Time Interaction ... 32
Supplementary table 19. PA Session Count Variable Age*Time Interaction ... 34
Supplementary table 20. PA Session Count Variable Gender*Time Interaction ... 36
Supplementary table 21. PA Session Count Variable Work From Home*Time Interaction ... 38
Supplementary table 22. PA Session Count Variable Living Alone*Time Interaction ... 40

**Supplementary Table 1.**

**Supplementary Table 1. Results of GEE Logistic Regression of meeting total WHO recommended levels of PA throughout April-January 2020 with Living Alone*Time Interaction**

| ***Independent Variable*** | ***Meeting WHO Recommended levels of MVPA & MSA*** | | | | | |
|---|---|---|---|---|---|---|
| | ***β*** | ***SE*** | ***OR*** | **95% CI** | **Wald $\chi 2$** | ***p-value*** |
| **Sociodemographic Factors** | | | | | | |
| **Gender (Female vs all others)** | -0.016 | 0.111 | 0.984 | (0.792-1.222) | 0.220 | 0.882 |
| **Age** | 0.000 | 0.004 | 1.000 | (0.992-1.009) | 0.002 | 0.960 |
| **Ethnicity (White vs all others)** | -0.096 | 0.176 | 0.909 | (0.644-1.283) | 0.295 | 0.587 |
| **Employment/Home Working (ref. Unemployed)** | | | | | 0.775 | 0.679 |
| *Employed + working from home* | -0.119 | 0.137 | 0.888 | (0.679-1.160) | | 0.382 |
| *Employed + attending workplace* | -0.068 | 0.160 | 0.934 | (0.682-1.279) | | 0.670 |
| **Socioeconomic Index (ref. Lowest)** | | | | | 11.112 | 0.011 |
| *Highest* | 0.859 | 0.359 | 2.362 | (1.169-4.772) | | 0.017 |
| *Upper* | 0.837 | 0.358 | 2.310 | (1.146-4.656) | | 0.019 |
| *Middle* | 0.527 | 0.364 | 1.693 | (0.829-3.458) | | 0.148 |
| **COVID-19 Situational Factors** | | | | | | |
| **Home Exercise Space Access** | -0.072 | 0.115 | 0.931 | (0.743-1.166) | 0.392 | 0.531 |
| **Living Alone** | -0.403 | 0.191 | 0.668 | (0.460-0.971) | 0.438 | 0.508 |
| **Living Alone*Time Interaction (ref. Baseline)** | | | | | 12.180 | 0.007 |
| Living Alone*1-month | 0.246 | 0.199 | | | | 0.217 |
| Living Alone*3-months | 0.723 | 0.219 | | | | 0.001 |
| Living Alone*6-months | 0.270 | 0.257 | | | | 0.293 |
| **Isolation (Strict vs all others)** | -0.583 | 0.198 | 0.558 | (0.379-0.823) | 8.658 | 0.003 |
| **Risk from COVID-19 (High vs all others)** | -0.178 | 0.078 | 0.837 | (0.719-0.974) | 5.271 | 0.022 |
| **Composite Quality of Life Index** | 0.250 | 0.054 | 1.285 | (1.155-1.428) | 21.474 | <0.001 |
| **Lifestyle & Health Factors** | | | | | | |

| | | | | | | |
|---|---|---|---|---|---|---|
| **BMI (Overweight vs all others)** | -0.414 | 0.093 | 0.661 | (0.550-0.794) | 19.693 | <0.001 |
| **Risk Alcohol Consumption (>14 weekly units)** | -0.188 | 0.102 | 0.828 | (0.679-1.011) | 3.444 | 0.063 |
| **Smoking (Smokers vs all others)** | -0.302 | 0.158 | 0.739 | (0.542-1.008) | 3.650 | 0.056 |
| **Chronic Health Condition** | -0.237 | 0.207 | 0.789 | (0.526-1.183) | 1.311 | 0.252 |
| **Suspected/Confirmed COVID-19 Infection** | -0.035 | 0.106 | 0.965 | (0.785-1.188) | 0.110 | 0.740 |
| **Meeting WHO Reccommended Levels (meeting neither):** | | | | | 319.502 | <0.001 |
| *MSA & MVPA before COVID-19* | 2.301 | 0.133 | 9.989 | (7.698-12.962) | | <0.001 |
| *MVPA only before COVID-19* | 0.790 | 0.141 | 2.203 | (1.672-2.902) | | <0.001 |
| *MSA only before COVID-19* | 1.309 | 0.149 | 3.704 | (2.764-4.964) | | <0.001 |
| **Time (Ref. Baseline)** | | | | | 11.442 | 0.010 |
| *1 month* | -0.186 | 0.781 | 0.830 | (0.712-0.967) | | 0.017 |
| *3 months* | -0.548 | 0.088 | 0.578 | (0.487-0.687) | | <0.001 |
| *6 months* | -0.559 | 0.098 | 0.571 | (0.472-0.692) | | <0.001 |

**Supplementary Table 2.**

**Supplementary Table 2. Results of GEE Logistic Regression of meeting total WHO recommended levels of PA throughout April-January 2020 with PA*Time Interaction**

| *Independent Variable* | *Meeting WHO Recommended levels of MVPA & MSA* | | | | | |
|---|---|---|---|---|---|---|
| *n=1908* | ***β*** | ***SE*** | ***OR*** | **95% CI** | **Wald $\chi 2$** | ***p-value*** |
| **Sociodemographic Factors** | | | | | | |
| **Gender (Female vs all others)** | -0.015 | 0.111 | 0.985 | (0.793-1.223) | 0.019 | 0.889 |
| **Age** | 0.000 | 0.004 | 1.000 | (0.992-1.009) | 0.005 | 0.942 |
| **Ethnicity (White vs all others)** | -0.083 | 0.176 | 0.921 | (0.652-1.300) | 0.219 | 0.639 |
| **Employment/Home Working (ref. Unemployed)** | | | | | 0.680 | 0.712 |
| *Employed + working from home* | -0.112 | 0.137 | 0.894 | (0.684-1.169) | | 0.413 |
| *Employed + attending workplace* | -0.065 | 0.161 | 0.937 | (0.684-1.283) | | 0.683 |
| **Socioeconomic Index (ref. Lowest)** | | | | | 11.273 | 0.010 |
| *Highest* | 0.895 | 0.360 | 2.448 | (1.208-4.961) | | 0.013 |
| *Upper* | 0.874 | 0.359 | 2.397 | (1.186-4.843) | | 0.015 |
| *Middle* | 0.571 | 0.366 | 1.770 | (0.865-3.623) | | 0.118 |
| **COVID-19 Situational Factors** | | | | | | |
| **Home Exercise Space Access** | -0.078 | 0.115 | 0.925 | (0.738-1.158) | 0.467 | 0.494 |
| **Living Alone** | -0.133 | 0.141 | 0.875 | (0.663-1.155) | 0.888 | 0.346 |
| **Isolation (Strict vs all others)** | -0.605 | 0.199 | 0.546 | (0.370-0.806) | 9.259 | 0.002 |
| **Risk from COVID-19 (High vs all others)** | -0.173 | 0.078 | 0.841 | (0.722-0.979) | 4.971 | 0.026 |
| **Composite Quality of Life Index** | 0.253 | 0.054 | 1.288 | (1.159-1.432) | 21.969 | <0.001 |
| **Lifestyle & Health Factors** | | | | | | |
| **BMI (Overweight vs all others)** | -0.421 | 0.093 | 0.657 | (0.547-0.789) | 20.245 | <0.001 |
| **Risk Alcohol Consumption (>14 weekly units)** | -0.185 | 0.102 | 0.831 | (0.681-1.015) | 3.290 | 0.070 |
| **Smoking (Smokers vs all others)** | -0.303 | 0.159 | 0.739 | (0.541-1.009) | 3.631 | 0.057 |
| **Chronic Health Condition** | -0.241 | 0.208 | 0.786 | (0.523-1.181) | 1.341 | 0.247 |

| | | | | | | |
|---|---|---|---|---|---|---|
| **Suspected/Confirmed COVID-19 Infection** | -0.025 | 0.105 | 0.975 | (0.794-1.198) | 0.056 | 0.813 |
| **Meeting WHO Reccommended Levels:** | | | | | 312.906 | <0.001 |
| *MSA & MVPA before COVID-19* | 2.086 | 0.169 | 8.051 | (5.776-11.222) | | <0.001 |
| *MVPA only before COVID-19* | 0.432 | 0.178 | 1.540 | (1.086-2.184) | | 0.015 |
| *MSA only before COVID-19* | 1.028 | 0.190 | 2.796 | (1.926-4.057) | | <0.001 |
| **Meeting WHO Reccommended Levels*Time Interaction Term** | | | | | 18.980 | 0.025 |
| *MSA & MVPA before COVID-19**1-month | 0.108 | 0.190 | | | | 0.570 |
| *MSA & MVPA before COVID-19**3-months | 0.372 | 0.215 | | | | 0.083 |
| *MSA & MVPA before COVID-19**6-months | 0.730 | 0.298 | | | | 0.015 |
| *MVPA only before COVID-19**1-month | 0.442 | 0.191 | | | | 0.021 |
| *MVPA only before COVID-19**3-months | 0.435 | 0.233 | | | | 0.062 |
| *MVPA only before COVID-19**6-months | 1.012 | 0.307 | | | | 0.001 |
| *MSA only before COVID-19**1-month | 0.066 | 0.223 | | | | 0.767 |
| *MSA only before COVID-19**3-months | 0.460 | 0.250 | | | | 0.066 |
| *MSA only before COVID-19**6-months | 0.937 | 0.322 | | | | 0.004 |
| **Time (Ref. Baseline)** | | | | | 43.574 | <0.001 |
| *1 month* | -0.314 | 0.140 | 0.730 | (0.556-0.960) | | 0.024 |
| *3 months* | -0.760 | 0.168 | 0.468 | (0.336-0.650) | | <0.001 |
| *6 months* | -1.232 | 0.256 | 0.292 | (0.177-0.481) | | <0.001 |

**Supplementary table 3.**

**Supplementary table 3. Results of GEE Logistic Regression of meeting WHO recommended levels of MVPA throughout April 2020-January 2021 with Employment/Home working*Time Interaction**

| ***Independent Variable*** | ***Meeting WHO Recommended levels of MVPA*** | | | | | |
|---|---|---|---|---|---|---|
| | ***β*** | ***SE*** | ***OR*** | | **Wald $\chi 2$** | ***p-value*** |
| **Sociodemographic Factors** | | | | | | |
| **Gender (Female vs all others)** | -0.206 | 0.856 | 0.814 | (0.688-0.963) | 5.784 | 0.016 |
| **Age** | 0.014 | 0.004 | 1.014 | (1.007-1.021) | 15.063 | <0.001 |
| **Ethnicity (White vs all others)** | 0.375 | 0.156 | 1.456 | (1.071-1.978) | 5.761 | 0.016 |
| **Employment/Home Working (ref. Unemployed)** | | | | | 0.182 | 0.913 |
| *Employed + working from home* | 0.080 | 0.135 | 1.203 | (0.878-1.647) | | 0.553 |
| *Employed + attending workplace* | 0.184 | 0.160 | 1.203 | (0.878-1.647) | | 0.250 |
| **Employment/Home Working*Time Interaction Term** | | | | | 15.579 | 0.016 |
| *Employed + working from home**1-month | 0.043 | 0.134 | | | | 0.749 |
| *Employed + working from home**3-months | 0.135 | 0.139 | | | | 0.333 |
| *Employed + working from home**6-months | -0.313 | 0.152 | | | | 0.039 |
| *Employed + attending workplace**1-month | -0.140 | 0.169 | | | | 0.936 |
| *Employed + attending workplace**3-months | -0.183 | 0.180 | | | | 0.309 |
| *Employed + attending workplace**6-months | -0.451 | 0.194 | | | | 0.020 |
| **Socioeconomic Index (ref. Lowest)** | | | | | 18.900 | <0.001 |
| *Highest* | 0.485 | 0.192 | 1.625 | (1.116-2.366) | | 0.011 |
| *Upper* | 0.355 | 0.184 | 1.426 | (0.995-2.045) | | 0.053 |
| *Middle* | 0.047 | 0.190 | 1.048 | (0.721-1.522) | | 0.806 |
| **COVID-19 Situational Factors** | | | | | | |
| **Home Exercise Space Access** | -0.221 | 0.091 | 0.802 | (0.671-0.959) | 5.882 | 0.015 |
| **Living Alone** | -0.071 | 0.113 | 0.932 | (0.747-1.162) | 0.393 | 0.531 |

| | | | | | | |
|---|---|---|---|---|---|---|
| **Isolation (Strict vs all others)** | -0.882 | 0.154 | 0.414 | (0.306-0.559) | 32.975 | <0.001 |
| **Risk from COVID-19 (High vs all others)** | -0.083 | 0.063 | 0.921 | (0.813-1.042) | 1.700 | 0.192 |
| **Composite Quality of Life Index** | 0.240 | 0.042 | 1.272 | (1.171-1.381) | 32.857 | <0.001 |
| **Lifestyle & Health Factors** | | | | | | |
| **BMI (Overweight vs all others)** | -0.296 | 0.071 | 0.744 | (0.647-0.855) | 17.241 | <0.001 |
| **Risk Alcohol Consumption (>14 weekly units)** | -0.125 | 0.075 | 0.883 | (0.762-1.023) | 2.758 | 0.097 |
| **Smoking (Smokers vs all others)** | -0.108 | 0.119 | 0.898 | (0.712-1.113) | 0.824 | 0.364 |
| **Chronic Health Condition** | -0.628 | 0.137 | 0.534 | (0.408-0.697) | 21.155 | <0.001 |
| **Suspected/Confirmed COVID-19 Infection** | -0.023 | 0.084 | 0.977 | (0.829-1.151) | 0.077 | 0.782 |
| **Meeting WHO Reccommended Levels (meeting neither):** | | | | | 400.629 | <0.001 |
| *MSA & MVPA before COVID-19* | 1.544 | 0.110 | 4.684 | (3.775-5.813) | | <0.001 |
| *MVPA only before COVID-19* | 1.657 | 0.096 | 5.243 | (4.343-6.329) | | <0.001 |
| *MSA only before COVID-19* | 0.238 | 0.119 | 1.268 | (1.005-1.601) | | 0.045 |
| **Time (Ref. Baseline)** | | | | | 36.834 | <0.001 |
| *1 month* | -0.136 | 0.107 | 0.873 | (0.708-1.077) | | 0.205 |
| *3 months* | -0.355 | 0.112 | 0.701 | (0.563-0.873) | | 0.001 |
| *6 months* | 0.092 | 0.117 | 0.912 | (0.726-1.147) | | 0.432 |

**Supplementary Table 4.**

**Supplementary Table 4. Results of GEE Logistic Regression of meeting WHO recommended levels of MSA throughout April-January 2021 with Gender*Time interaction only**

| *Independent Variable* | *Meeting WHO Recommended levels of MSA* | | | | | |
|---|---|---|---|---|---|---|
| | *β* | *SE* | *OR* | | **Wald $\chi 2$** | *p-value* |
| **Sociodemographic Factors** | | | | | | |
| **Gender (Female vs all others)** | 0.134 | 0.124 | 1.144 | (0.898-1.458) | 1.162 | 0.281 |
| **Gender*Time Interaction Term** | | | | | 10.617 | 0.014 |
| *Gender (Female)**1-month | -0.184 | 0.124 | | | | 0.138 |
| *Gender (Female)*3*-months | -0.436 | 0.137 | | | | 0.001 |
| *Gender (Female)*6*-months | -0.339 | 0.148 | | | | 0.022 |
| **Age** | -0.009 | 0.004 | 0.991 | (0.984-0.998) | 6.128 | 0.013 |
| **Ethnicity (White vs all others)** | -0.415 | 0.198 | 0.660 | (0.448-0.974) | 4.387 | 0.036 |
| **Employment/Home Working (ref. Unemployed)** | | | | | 1.703 | 0.427 |
| *Employed + working from home* | -0.120 | 0.120 | 0.988 | (0.780-1.250) | | 0.921 |
| *Employed + attending workplace* | -0.157 | 0.140 | 0.855 | (0.650-1.124) | | 0.262 |
| **Socioeconomic Index (ref. Lowest)** | | | | | 1.867 | 0.601 |
| *Highest* | 0.366 | 0.319 | 1.442 | (0.773-2.693) | | 0.250 |
| *Upper* | 0.324 | 0.315 | 1.383 | (0.746-2.563) | | 0.303 |
| *Middle* | 0.254 | 0.319 | 1.289 | (0.690-2.408) | | 0.427 |
| **COVID-19 Situational Factors** | | | | | | |
| **Home Exercise Space Access** | -0.063 | 0.102 | 0.939 | (0.769-1.145) | 0.389 | 0.533 |
| **Living Alone** | -0.181 | 0.125 | 0.835 | (0.653-1.067) | 2.078 | 0.149 |

| | | | | | | |
|---|---|---|---|---|---|---|
| **Isolation (Strict vs all others)** | -0.034 | 0.148 | 0.966 | (0.723-1.290) | 0.054 | 0.816 |
| **Risk from COVID-19 (High vs all others)** | -0.011 | 0.065 | 0.989 | (0.870-1.125) | 0.027 | 0.869 |
| **Composite Quality of Life Index** | 0.249 | 0.046 | 1.283 | (1.174-1.403) | 30.098 | <0.001 |
| **Lifestyle & Health Factors** | | | | | | |
| **BMI (Overweight vs all others)** | -0.367 | 0.077 | 0.693 | (0.596-0.806) | 22.631 | <0.001 |
| **Risk Alcohol Consumption (>14 weekly units)** | -0.243 | 0.077 | 0.785 | (0.674-0.913) | 9.830 | <0.001 |
| **Smoking (Smokers vs all others)** | -0.269 | 0.129 | 0.764 | (0.594-0.958) | 4.339 | 0.037 |
| **Chronic Health Condition** | -0.131 | 0.160 | 0.877 | (0.642-1.200) | 0.669 | 0.413 |
| **Suspected/Confirmed COVID-19 Infection** | -0.046 | 0.088 | 0.955 | (0.804-1.134) | 0.278 | 0.598 |
| **Meeting WHO Reccommended Levels (meeting neither):** | | | | | 502.930 | <0.001 |
| *MSA & MVPA before COVID-19* | 2.035 | 0.117 | 7.650 | (6.078-9.628) | | <0.001 |
| *MVPA only before COVID-19* | 0.073 | 0.116 | 1.075 | (0.857-1.350) | | 0.531 |
| *MSA only before COVID-19* | 1.968 | 0.123 | 7.156 | (5.623-9.106) | | <0.001 |
| **Time (Ref. Baseline)** | | | | | 15.563 | <0.001 |
| *1 month* | 0.071 | 0.104 | 1.073 | (0.875-1.317) | | 0.497 |
| *3 months* | 0.043 | 0.114 | 1.044 | (0.834-1.306) | | 0.709 |
| *6 months* | -0.091 | 0.121 | 0.913 | (0.719-1.158) | | 0.451 |

**Supplementary Table 5.**

**Supplementary table 5. Results of GEE Logistic Regression of meeting WHO recommended levels of MSA throughout April 2020-January 2021 with Age*Time Interaction**

| *Independent Variable* | *Meeting WHO Recommended levels of MSA* | | | | | |
|---|---|---|---|---|---|---|
| | ***β*** | ***SE*** | ***OR*** | | **Wald $\chi 2$** | ***p-value*** |
| **Sociodemographic Factors** | | | | | | |
| **Gender (Female vs all others)** | -0.073 | 0.097 | 0.929 | (0.768-1.124) | 0.569 | 0.451 |
| **Age** | -0.017 | 0.004 | 0.983 | (0.975-0.992) | 5.085 | 0.024 |
| **Age*Time Interaction Term** | | | | | 11.785 | 0.008 |
| *Age**1-month | 0.006 | 0.004 | | | | 0.122 |
| *Age*3*-months | 0.012 | 0.004 | | | | 0.008 |
| *Age*6*-months | 0.015 | 0.005 | | | | 0.001 |
| **Ethnicity (White vs all others)** | -0.415 | 0.198 | 0.661 | (0.449-0.973) | 4.408 | 0.036 |
| **Employment/Home Working (ref. Unemployed)** | | | | | 1.790 | 0.409 |
| *Employed + working from home* | -0.015 | 0.120 | 0.985 | (0.778-1.246) | | 0.899 |
| *Employed + attending workplace* | -0.162 | 0.140 | 0.850 | (0.647-1.118) | | 0.246 |
| **Socioeconomic Index (ref. Lowest)** | | | | | 1.681 | 0.641 |
| *Highest* | 0.355 | 0.319 | 1.426 | (0.762-2.666) | | 0.267 |
| *Upper* | 0.312 | 0.316 | 1.366 | (0.736-2.535) | | 0.323 |
| *Middle* | 0.250 | 0.320 | 1.284 | (0.686-2.402) | | 0.435 |
| **COVID-19 Situational Factors** | | | | | | |
| **Home Exercise Space Access** | -0.061 | 0.102 | 0.940 | (0.771-1.148) | 0.366 | 0.545 |
| **Living Alone** | -0.180 | 0.125 | 0.836 | (0.654-1.068) | 2.060 | 0.151 |
| **Isolation (Strict vs all others)** | -0.008 | 0.147 | 0.992 | (0.743-1.323) | 0.003 | 0.955 |
| **Risk from COVID-19 (High vs all others)** | -0.010 | 0.066 | 0.992 | (0.871-1.126) | 0.239 | 0.625 |

| | | | | | | |
|---|---|---|---|---|---|---|
| **Composite Quality of Life Index** | 0.249 | 0.045 | 1.283 | (1.174-1.402) | 30.094 | <0.001 |
| **Lifestyle & Health Factors** | | | | | | |
| **BMI (Overweight vs all others)** | -0.365 | 0.077 | 0.695 | (0.597-0.808) | 22.381 | <0.001 |
| **Risk Alcohol Consumption (>14 weekly units)** | -0.225 | 0.077 | 0.798 | (0.686-0.929) | 8.465 | 0.004 |
| **Smoking (Smokers vs all others)** | -0.290 | 0.130 | 0.748 | (0.58-0.964) | 5.014 | 0.025 |
| **Chronic Health Condition** | -0.133 | 0.160 | 0.876 | (0.640-1.197) | 0.693 | 0.405 |
| **Suspected/Confirmed COVID-19 Infection** | -0.043 | 0.088 | 0.958 | (0.806-1.138) | 0.239 | 0.625 |
| **Meeting WHO Reccommended Levels (meeting neither):** | | | | | 503.658 | <0.001 |
| *MSA & MVPA before COVID-19* | 2.037 | 0.117 | 7.664 | (6.090-9.645) | | <0.001 |
| *MVPA only before COVID-19* | 0.073 | 0.116 | 1.076 | (0.857-1.352) | | 0.528 |
| *MSA only before COVID-19* | 1.971 | 0.123 | 7.175 | (5.637-9.132) | | <0.001 |
| **Time (Ref. Baseline)** | | | | | 21.638 | <0.001 |
| *1 month* | -0.366 | 0.212 | 0.693 | (0.458-1.050) | | 0.084 |
| *3 months* | -0.839 | 0.232 | 0.432 | (0.275-0.681) | | <0.001 |
| *6 months* | -1.102 | 0.256 | 0.332 | (0.201-0.549) | | <0.001 |

**Supplementary table 6 (Pairwise Comparisons of MVPA & MSA (total sample)).**

##### Estimates

| Time | Mean | Std. Error | 95% Wald Confidence Interval Lower | 95% Wald Confidence Interval Upper |
|---|---|---|---|---|
| 7 | .14 | .008 | .13 | .16 |
| 4 | .15 | .008 | .14 | .17 |
| 2 | .18 | .009 | .16 | .20 |
| 1 | .20 | .009 | .18 | .21 |

##### Pairwise Comparisons

| (I) Time | (J) Time | Mean Difference (I-J) | Std. Error | df | Sequential Sidak Sig. | 95% Wald Confidence Interval for Difference[a] Lower | 95% Wald Confidence Interval for Difference[a] Upper |
|---|---|---|---|---|---|---|---|
| 7 | 4 | -.01 | .009 | 1 | .244 | -.03 | .01 |
| | 2 | -.04[b] | .009 | 1 | .001 | -.06 | -.01 |
| | 1 | -.05[b] | .010 | 1 | .000 | -.08 | -.03 |
| 4 | 7 | .01 | .009 | 1 | .244 | -.01 | .03 |
| | 2 | -.03[b] | .009 | 1 | .012 | -.05 | .00 |
| | 1 | -.04[b] | .009 | 1 | .000 | -.07 | -.02 |
| 2 | 7 | .04[b] | .009 | 1 | .001 | .01 | .06 |
| | 4 | .03[b] | .009 | 1 | .012 | .00 | .05 |
| | 1 | -.02 | .009 | 1 | .077 | -.04 | .00 |
| 1 | 7 | .05[b] | .010 | 1 | .000 | .03 | .08 |
| | 4 | .04[b] | .009 | 1 | .000 | .02 | .07 |
| | 2 | .02 | .009 | 1 | .077 | .00 | .04 |

Pairwise comparisons of estimated marginal means based on the original scale of dependent variable Meeting total WHO recommended PA 2 levels with 1 = meeting both

a. Confidence interval bounds are approximate.

b. The mean difference is significant at the .05 level.

**Supplementary table 7 (Pairwise Comparisons of MVPA (total sample)).**

**Estimates**

| Time | Mean | Std. Error | 95% Wald Confidence Interval Lower | Upper |
|---|---|---|---|---|
| 7 | .39 | .012 | .37 | .41 |
| 4 | .40 | .011 | .37 | .42 |
| 2 | .42 | .011 | .40 | .45 |
| 1 | .44 | .011 | .42 | .46 |

**Pairwise Comparisons**

| (I) Time | (J) Time | Mean Difference (I-J) | Std. Error | df | Sequential Sidak Sig. | 95% Wald Confidence Interval for Difference[a] Lower | Upper |
|---|---|---|---|---|---|---|---|
| 7 | 4 | -.01 | .012 | 1 | .605 | -.03 | .02 |
| | 2 | -.03[b] | .012 | 1 | .016 | -.06 | .00 |
| | 1 | -.05[b] | .013 | 1 | .001 | -.08 | -.01 |
| 4 | 7 | .01 | .012 | 1 | .605 | -.02 | .03 |
| | 2 | -.03[b] | .011 | 1 | .031 | -.05 | .00 |
| | 1 | -.04[b] | .012 | 1 | .002 | -.07 | -.01 |
| 2 | 7 | .03[b] | .012 | 1 | .016 | .00 | .06 |
| | 4 | .03[b] | .011 | 1 | .031 | .00 | .05 |
| | 1 | -.01 | .011 | 1 | .380 | -.04 | .01 |
| 1 | 7 | .05[b] | .013 | 1 | .001 | .01 | .08 |
| | 4 | .04[b] | .012 | 1 | .002 | .01 | .07 |
| | 2 | .01 | .011 | 1 | .380 | -.01 | .04 |

Pairwise comparisons of estimated marginal means based on the original scale of dependent variable meeting_mvpaSIN

a. Confidence interval bounds are approximate.

b. The mean difference is significant at the .05 level.

**Supplementary table 8 (Pairwise Comparisons of MSA (total sample)).**

**Estimates**

| Time | Mean | Std. Error | 95% Wald Confidence Interval | |
|---|---|---|---|---|
| | | | Lower | Upper |
| 7 | .29 | .011 | .27 | .31 |
| 4 | .31 | .010 | .29 | .33 |
| 2 | .33 | .011 | .31 | .35 |
| 1 | .34 | .011 | .32 | .36 |

**Pairwise Comparisons**

| (I) Time | (J) Time | Mean Difference (I-J) | Std. Error | df | Sequential Sidak Sig. | 95% Wald Confidence Interval for Difference[a] | |
|---|---|---|---|---|---|---|---|
| | | | | | | Lower | Upper |
| 7 | 4 | -.02 | .010 | 1 | .183 | -.04 | .01 |
| | 2 | -.04[b] | .011 | 1 | .001 | -.07 | -.01 |
| | 1 | -.05[b] | .011 | 1 | .000 | -.08 | -.02 |
| 4 | 7 | .02 | .010 | 1 | .183 | -.01 | .04 |
| | 2 | -.03[b] | .010 | 1 | .026 | -.05 | .00 |
| | 1 | -.04[b] | .010 | 1 | .002 | -.06 | -.01 |
| 2 | 7 | .04[b] | .011 | 1 | .001 | .01 | .07 |
| | 4 | .03[b] | .010 | 1 | .026 | .00 | .05 |
| | 1 | -.01 | .009 | 1 | .210 | -.03 | .01 |
| 1 | 7 | .05[b] | .011 | 1 | .000 | .02 | .08 |
| | 4 | .04[b] | .010 | 1 | .002 | .01 | .06 |
| | 2 | .01 | .009 | 1 | .210 | -.01 | .03 |

Pairwise comparisons of estimated marginal means based on the original scale of dependent variable meeting_msaSIN

a. Confidence interval bounds are approximate.

b. The mean difference is significant at the .05 level.

**Supplementary table 9 (Complete Case Pairwise comparisons of MVPA & MSA).**

**Estimates**

| Time | Mean | Std. Error | 95% Wald Confidence Interval Lower | Upper |
|---|---|---|---|---|
| 7 | .14 | .009 | .13 | .16 |
| 4 | .16 | .009 | .14 | .18 |
| 2 | .18 | .010 | .16 | .20 |
| 1 | .20 | .010 | .18 | .22 |

**Pairwise Comparisons**

| (I) Time | (J) Time | Mean Difference (I-J) | Std. Error | df | Sequential Sidak Sig. | 95% Wald Confidence Interval for Difference[a] Lower | Upper |
|---|---|---|---|---|---|---|---|
| 7 | 4 | -.01 | .012 | 1 | .605 | -.03 | .02 |
| | 2 | -.03[b] | .012 | 1 | .016 | -.06 | .00 |
| | 1 | -.05[b] | .013 | 1 | .001 | -.08 | -.01 |
| 4 | 7 | .01 | .012 | 1 | .605 | -.02 | .03 |
| | 2 | -.03[b] | .011 | 1 | .031 | -.05 | .00 |
| | 1 | -.04[b] | .012 | 1 | .002 | -.07 | -.01 |
| 2 | 7 | .03[b] | .012 | 1 | .016 | .00 | .06 |
| | 4 | .03[b] | .011 | 1 | .031 | .00 | .05 |
| | 1 | -.01 | .011 | 1 | .380 | -.04 | .01 |
| 1 | 7 | .05[b] | .013 | 1 | .001 | .01 | .08 |
| | 4 | .04[b] | .012 | 1 | .002 | .01 | .07 |
| | 2 | .01 | .011 | 1 | .380 | -.01 | .04 |

Pairwise comparisons of estimated marginal means based on the original scale of dependent variable meeting_mvpaSIN

a. Confidence interval bounds are approximate.

b. The mean difference is significant at the .05 level.

**Supplementary table 10 (Complete Case Pairwise comparisons of MVPA).**

**Estimates**

| Time | Mean | Std. Error | 95% Wald Confidence Interval Lower | 95% Wald Confidence Interval Upper |
|---|---|---|---|---|
| 7 | .29 | .012 | .27 | .32 |
| 4 | .31 | .012 | .29 | .34 |
| 2 | .34 | .012 | .31 | .36 |
| 1 | .34 | .012 | .32 | .36 |

**Pairwise Comparisons**

| (I) Time | (J) Time | Mean Difference (I-J) | Std. Error | df | Sequential Sidak Sig. | 95% Wald Confidence Interval for Difference[a] Lower | 95% Wald Confidence Interval for Difference[a] Upper |
|---|---|---|---|---|---|---|---|
| 7 | 4 | -.02 | .010 | 1 | .098 | -.05 | .00 |
| | 2 | -.04[b] | .011 | 1 | .001 | -.07 | -.01 |
| | 1 | -.05[b] | .012 | 1 | .000 | -.08 | -.02 |
| 4 | 7 | .02 | .010 | 1 | .098 | .00 | .05 |
| | 2 | -.02 | .011 | 1 | .098 | -.05 | .00 |
| | 1 | -.03 | .011 | 1 | .083 | -.05 | .00 |
| 2 | 7 | .04[b] | .011 | 1 | .001 | .01 | .07 |
| | 4 | .02 | .011 | 1 | .098 | .00 | .05 |
| | 1 | .00 | .010 | 1 | .695 | -.02 | .02 |
| 1 | 7 | .05[b] | .012 | 1 | .000 | .02 | .08 |
| | 4 | .03 | .011 | 1 | .083 | .00 | .05 |
| | 2 | .00 | .010 | 1 | .695 | -.02 | .02 |

Pairwise comparisons of estimated marginal means based on the original scale of dependent variable meeting_msaSIN

a. Confidence interval bounds are approximate.

b. The mean difference is significant at the .05 level.

**Supplementary table 11 (Complete Case Pairwise comparisons of MSA).**

**Estimates**

| Time | Mean | Std. Error | 95% Wald Confidence Interval | |
|---|---|---|---|---|
| | | | Lower | Upper |
| 7 | .40 | .012 | .37 | .42 |
| 4 | .40 | .012 | .38 | .43 |
| 2 | .44 | .013 | .41 | .46 |
| 1 | .45 | .013 | .43 | .48 |

**Pairwise Comparisons**

| (I) Time | (J) Time | Mean Difference (I-J) | Std. Error | df | Sequential Sidak Sig. | 95% Wald Confidence Interval for Difference[a] | |
|---|---|---|---|---|---|---|---|
| | | | | | | Lower | Upper |
| 7 | 4 | -.01 | .012 | 1 | .602 | -.03 | .02 |
| | 2 | -.04[b] | .012 | 1 | .003 | -.07 | -.01 |
| | 1 | -.06[b] | .014 | 1 | .000 | -.09 | -.02 |
| 4 | 7 | .01 | .012 | 1 | .602 | -.02 | .03 |
| | 2 | -.04[b] | .012 | 1 | .011 | -.06 | -.01 |
| | 1 | -.05[b] | .013 | 1 | .001 | -.08 | -.02 |
| 2 | 7 | .04[b] | .012 | 1 | .003 | .01 | .07 |
| | 4 | .04[b] | .012 | 1 | .011 | .01 | .06 |
| | 1 | -.01 | .012 | 1 | .413 | -.04 | .01 |
| 1 | 7 | .06[b] | .014 | 1 | .000 | .02 | .09 |
| | 4 | .05[b] | .013 | 1 | .001 | .02 | .08 |
| | 2 | .01 | .012 | 1 | .413 | -.01 | .04 |

Pairwise comparisons of estimated marginal means based on the original scale of dependent variable meeting_mvpaSIN

a. Confidence interval bounds are approximate.

b. The mean difference is significant at the .05 level.

**Supplementary table 12 (MSA & MVPA with both time interactions).**

**Table 3. Results of GEE Logistic Regression of meeting total WHO recommended levels of PA throughout April-January 2020.**

| *Independent Variable* | *Meeting WHO Recommended levels of MVPA & MSA* | | | | | |
|---|---|---|---|---|---|---|
| | ***β*** | ***SE*** | ***OR*** | **95% CI** | **Wald $\chi 2$** | ***p-value*** |
| **Sociodemographic Factors** | | | | | | |
| **Gender (Female vs all others)** | -0.021 | 0.110 | 0.979 | (0.790-1.214) | 0.037 | 0.847 |
| **Age** | 0.000 | 0.004 | 1.000 | (0.992-1.009) | 0.011 | 0.916 |
| **Ethnicity (White vs all others)** | -0.067 | 0.176 | 0.953 | (0.663-1.320) | 0.144 | 0.704 |
| **Employment/Home Working (ref. Unemployed)** | | | | | 0.446 | 0.800 |
| *Employed + working from home* | -0.091 | 0.136 | 0.913 | (0.700-1.192) | | 0.466 |
| *Employed + attending workplace* | -0.061 | 0.161 | 0.941 | (0.688-1.288) | | 0.143 |
| **Socioeconomic Index (ref. Lowest)** | | | | | 10.655 | 0.014 |
| *Highest* | 0.843 | 0.358 | 2.323 | (1.151-4.688) | | 0.019 |
| *Upper* | 0.831 | 0.357 | 2.296 | (1.141-4.4621) | | 0.020 |
| *Middle* | 0.527 | 0.364 | 1.694 | (0.831-3.457) | | 0.147 |
| **COVID-19 Situational Factors** | | | | | | |
| **Home Exercise Space Access** | -0.051 | 0.115 | 0.950 | (0.759-1.190) | 0.197 | 0.657 |
| **Living Alone** | -0.416 | 0.189 | 0.659 | (0.455-0.956) | 0.355 | 0.551 |
| **Isolation (Strict vs all others)** | -0.587 | 0.198 | 0.556 | (0.377-0.819) | 8.819 | 0.003 |
| **Risk from COVID-19 (High vs all others)** | -0.158 | 0.077 | 0.854 | (0.734-0.994) | 4.165 | 0.041 |
| **Composite Quality of Life Index** | 0.258 | 0.054 | 1.295 | (1.165-1.439) | 23.028 | <0.001 |
| **Lifestyle & Health Factors** | | | | | | |
| **BMI (Overweight vs all others)** | -0.429 | 0.093 | 0.651 | (0.543-0.781) | 21.404 | <0.001 |
| **Risk Alcohol Consumption (>14 weekly units)** | -0.192 | 0.101 | 0.825 | (0.676-1.006) | 3.600 | 0.058 |
| **Smoking (Smokers vs all others)** | -0.301 | 0.157 | 0.740 | (0.544-1.008) | 3.652 | 0.056 |
| **Chronic Health Condition** | -0.237 | 0.207 | 0.789 | (0.525-1.184) | 1.309 | 0.252 |

| | | | | | | |
|---|---|---|---|---|---|---|
| **Suspected/Confirmed COVID-19 Infection** | -0.003 | 0.104 | 0.997 | (0.812-1.222) | 0.001 | 0.973 |
| **Meeting WHO Reccommended Levels:** | | | | | 317.833 | <0.001 |
| *MSA & MVPA before COVID-19* | 2.106 | 0.170 | 8.218 | (5.895-11.457) | | <0.001 |
| *MVPA only before COVID-19* | 0.451 | 0.177 | 1.570 | (1.109-2.222) | | 0.011 |
| *MSA only before COVID-19* | 1.038 | 0.190 | 2.822 | (1.947-4.092) | | <0.001 |
| **Meeting WHO Reccommended Levels*Time Interaction Term** | | | | | 17.828 | 0.037 |
| *MSA & MVPA before COVID-19**1-month | 0.107 | 0.191 | | | | 0.576 |
| *MSA & MVPA before COVID-19**3-months | 0.361 | 0.214 | | | | 0.092 |
| *MSA & MVPA before COVID-19**6-months | 0.716 | 0.297 | | | | 0.016 |
| *MVPA only before COVID-19**1-month | 0.416 | 0.192 | | | | 0.030 |
| *MVPA only before COVID-19**3-months | 0.424 | 0.233 | | | | 0.069 |
| *MVPA only before COVID-19**6-months | 0.984 | 0.306 | | | | 0.001 |
| *MSA only before COVID-19**1-month | 0.072 | 0.220 | | | | 0.745 |
| *MSA only before COVID-19**3-months | 0.469 | 0.249 | | | | 0.060 |
| *MSA only before COVID-19**6-months | 0.934 | 0.321 | | | | 0.004 |
| **Time (Ref. Baseline)** | | | | | 11.714 | 0.008 |
| *1 month* | -0.343 | 0.142 | 0.709 | (0.537-0.937) | | 0.016 |
| *3 months* | -0.874 | 0.172 | 0.417 | (0.298-0.585) | | <0.001 |
| *6 months* | -1.272 | 0.256 | 0.280 | (0.170-0.463) | | <0.001 |

| | | | | | | |
|---|---|---|---|---|---|---|
| **Living Alone*Time Interaction (ref. Baseline)** | | | | | 12.253 | 0.007 |
| Living Alone*1-month | 0.260 | 0.198 | | | | 0.188 |
| Living Alone*3-months | 0.746 | 0.221 | | | | 0.001 |
| Living Alone*6-months | 0.322 | 0.256 | | | | 0.208 |

**Supplementary table 13 (MSA & MVPA with both time interactions).**

**Supplementary table 14. Results of GEE Logistic Regression of meeting WHO recommended levels of MSA throughout April 2020-January 2021 with Age*Time & Gender*Time interaction term.**

| *Independent Variable* | *Meeting WHO Recommended levels of MSA* | | | | | |
|---|---|---|---|---|---|---|
| | ***β*** | ***SE*** | ***OR*** | | **Wald $\chi2$** | ***p-value*** |
| **Sociodemographic Factors** | | | | | | |
| **Gender (Female vs all others)** | 0.123 | 0.124 | 1.131 | (0.887-1.442) | 1.238 | 0.226 |
| **Gender*Time Interaction Term** | | | | | 10.020 | 0.018 |
| *Gender (Female)**1-month | -0.173 | 0.124 | | | | 0.162 |
| *Gender (Female)*3*-months | -0.424 | 0.137 | | | | 0.002 |
| *Gender (Female)*6*-months | -0.326 | 0.148 | | | | 0.027 |
| **Age** | -0.016 | 0.004 | 0.984 | (0.975-0.992) | 5.296 | 0.021 |
| **Age*Time Interaction Term** | | | | | 10.020 | 0.012 |
| *Age**1-month | 0.006 | 0.004 | | | | 0.155 |
| *Age*3*-months | 0.011 | 0.004 | | | | 0.013 |
| *Age*6*-months | 0.015 | 0.005 | | | | 0.002 |
| **Ethnicity (White vs all others)** | -0.414 | 0.197 | 0.661 | (0.449-0.973) | 4.409 | 0.036 |
| **Employment/Home Working (ref. Unemployed)** | | | | | 1.728 | 0.421 |
| *Employed + working from home* | 0.000 | 0.120 | 1.000 | (0.791-1.264) | | 0.999 |
| *Employed + attending workplace* | -0.149 | 0.139 | 0.862 | (0.657-1.131) | | 0.284 |
| **Socioeconomic Index (ref. Lowest)** | | | | | 1.936 | 0.586 |
| *Highest* | 0.365 | 0.318 | 1.440 | (0.772-2.687) | | 0.251 |
| *Upper* | 0.331 | 0.314 | 1.392 | (0.752-2.578) | | 0.292 |
| *Middle* | 0.251 | 0.319 | 1.285 | (0.688-2.399) | | 0.432 |
| **COVID-19 Situational Factors** | | | | | | |

| | | | | | | |
|---|---|---|---|---|---|---|
| **Home Exercise Space Access** | -0.047 | 0.101 | 0.954 | (0.783-1.163) | 0.215 | 0.643 |
| **Living Alone** | -0.177 | 0.125 | 0.838 | (0.656-1.069) | 2.022 | 0.155 |
| **Isolation (Strict vs all others)** | -0.023 | 0.148 | 0.978 | (0.732-1.306) | 0.024 | 0.878 |
| **Risk from COVID-19 (High vs all others)** | -0.001 | 0.066 | 0.999 | (0.879-1.136) | 0.000 | 0.989 |
| **Composite Quality of Life Index** | 0.251 | 0.045 | 1.286 | (1.176-1.405) | 30.636 | <0.001 |
| **Lifestyle & Health Factors** | | | | | | |
| **BMI (Overweight vs all others)** | -0.375 | 0.077 | 0.688 | (0.591-0.799) | 23.795 | <0.001 |
| **Risk Alcohol Consumption (>14 weekly units)** | -0.241 | 0.773 | 0.786 | (0.676-0.915) | 9.671 | 0.002 |
| **Smoking (Smokers vs all others)** | -0.264 | 0.128 | 0.768 | (0.597-0.987) | 4.264 | 0.039 |
| **Chronic Health Condition** | -0.137 | 0.160 | 0.872 | (0.637-1.192) | 0.737 | 0.390 |
| **Suspected/Confirmed COVID-19 Infection** | -0.023 | 0.087 | 0.977 | (0.824-1.159) | 0.069 | 0.792 |
| **Meeting WHO Reccommended Levels (meeting neither):** | | | | | 513.880 | <0.001 |
| *MSA & MVPA before COVID-19* | 2.056 | 0.117 | 7.817 | (6.214-9.833) | | <0.001 |
| *MVPA only before COVID-19* | 0.078 | 0.116 | 1.081 | (0.862-1.356) | | 0.501 |
| *MSA only before COVID-19* | 1.984 | 0.123 | 7.274 | (5.720-9.250) | | <0.001 |
| **Time (Ref. Baseline)** | | | | | 17.190 | 0.001 |
| *1 month* | -0.366 | 0.212 | 0.693 | (0.458-1.050) | | 0.349 |
| *3 months* | -0.498 | 0.251 | 0.607 | (0.371-0.994) | | 0.047 |
| *6 months* | -0.210 | 0.224 | 0.810 | (0.522-1.258) | | 0.003 |

**Supplementary table 14 (Complete Case Models – Meeting WHO Total Guidelines)**

Tests of Model Effects

| Source | Type III Wald Chi-Square | df | Sig. |
|---|---|---|---|
| (Intercept) | 102.673 | 1 | 0 |
| Time | 11.714 | 3 | 0.008 |
| gender 2 levels including PNS | 0.037 | 1 | 0.847 |
| perceived risk at BL including DN | 4.165 | 1 | 0.041 |
| Composite_QoL | 23.028 | 1 | 0 |
| age_continuous | 0.011 | 1 | 0.916 |
| Composite occupation and working from home | 0.446 | 2 | 0.8 |
| Binary Living With Others (Alone=1 With Others=0). | 0.355 | 1 | 0.551 |
| Access to a garden Binary Var | 0.197 | 1 | 0.657 |
| Binary Isolation (Total Isolation=1 All Else = 0). | 8.819 | 1 | 0.003 |
| ethnicity dichotomised including PNS | 0.144 | 1 | 0.704 |
| Composite Housing/Income(0-2) + Education (0-1) Score | 10.655 | 3 | 0.014 |
| Confirmed and suspected Covid-19 2 levels | 0.001 | 1 | 0.973 |
| Limiting_Condition | 1.309 | 1 | 0.252 |
| BMI 2 levels including PNTS and DN | 21.404 | 1 | 0 |
| Meeting_BEF_categorical | 317.833 | 3 | 0 |
| Alcohol_Risk | 3.6 | 1 | 0.058 |
| Smoking baseline binary yes or no | 3.652 | 1 | 0.056 |
| Time * Binary Living With Others (Alone=1 With Others=0). | 12.253 | 3 | 0.007 |
| Time * Meeting_BEF_categorical | 17.828 | 9 | 0.037 |

Dependent Variable: Meeting total WHO recommended PA 2 levels with 1 = meeting both

| Parameter | B | Std. Error | 95% Wald Confidenc | | Hypothesis Test | | | Exp(B) | 95% Wald Confidenc | |
|---|---|---|---|---|---|---|---|---|---|---|
| | | | Lower | Upper | Wald Chi- | df | Sig. | | Lower | Upper |
| (Intercept) | -3.232 | 0.5043 | -4.221 | -2.244 | 41.084 | 1 | 0 | 0.039 | 0.015 | 0.106 |
| [Time=7] | -1.272 | 0.256 | -1.774 | -0.77 | 24.696 | 1 | 0 | 0.28 | 0.17 | 0.463 |
| [Time=4] | -0.874 | 0.1724 | -1.212 | -0.536 | 25.682 | 1 | 0 | 0.417 | 0.298 | 0.585 |
| [Time=2] | -0.343 | 0.142 | -0.622 | -0.065 | 5.851 | 1 | 0.016 | 0.709 | 0.537 | 0.937 |
| [Time=1] | 0a | . | . | . | . | . | . | 1 | . | . |
| [gender 2 levels including PNS=1.00] | -0.021 | 0.1098 | -0.236 | 0.194 | 0.037 | 1 | 0.847 | 0.979 | 0.79 | 1.214 |
| [gender 2 levels including PNS=.00] | 0a | . | . | . | . | . | . | 1 | . | . |
| [perceived risk at BL including DN=1.00] | -0.158 | 0.0774 | -0.31 | -0.006 | 4.165 | 1 | 0.041 | 0.854 | 0.734 | 0.994 |
| [perceived risk at BL including DN=.00] | 0a | . | . | . | . | . | . | 1 | . | . |
| Composite_QoL | 0.258 | 0.0538 | 0.153 | 0.364 | 23.028 | 1 | 0 | 1.295 | 1.165 | 1.439 |
| age_continuous | 0 | 0.0042 | -0.008 | 0.009 | 0.011 | 1 | 0.916 | 1 | 0.992 | 1.009 |
| [Composite occupation and working from home=3.00] | -0.061 | 0.1601 | -0.374 | 0.253 | 0.143 | 1 | 0.705 | 0.941 | 0.688 | 1.288 |
| [Composite occupation and working from home=2.00] | -0.091 | 0.1359 | -0.357 | 0.176 | 0.446 | 1 | 0.504 | 0.913 | 0.7 | 1.192 |
| [Composite occupation and working from home=1.00] | 0a | . | . | . | . | . | . | 1 | . | . |
| [Binary Living With Others (Alone=1 With Others=0).=1.00] | -0.416 | 0.1893 | -0.787 | -0.045 | 4.835 | 1 | 0.028 | 0.659 | 0.455 | 0.956 |
| [Binary Living With Others (Alone=1 With Others=0).=.00] | 0a | . | . | . | . | . | . | 1 | . | . |
| [Access to a garden Binary Var=1.00] | -0.051 | 0.1149 | -0.276 | 0.174 | 0.197 | 1 | 0.657 | 0.95 | 0.759 | 1.19 |
| [Access to a garden Binary Var=.00] | 0a | . | . | . | . | . | . | 1 | . | . |
| [Binary Isolation (Total Isolation=1 All Else = 0).=1.00] | -0.587 | 0.1978 | -0.975 | -0.2 | 8.819 | 1 | 0.003 | 0.556 | 0.377 | 0.819 |
| [Binary Isolation (Total Isolation=1 All Else = 0).=.00] | 0a | . | . | . | . | . | . | 1 | . | . |
| [ethnicity dichotomised including PNS=1.00] | -0.067 | 0.1757 | -0.411 | 0.278 | 0.144 | 1 | 0.704 | 0.935 | 0.663 | 1.32 |
| [ethnicity dichotomised including PNS=.00] | 0a | . | . | . | . | . | . | 1 | . | . |
| [Composite Housing/Income(0-2) + Education (0-1) Score=3.00] | 0.843 | 0.3583 | 0.14 | 1.545 | 5.532 | 1 | 0.019 | 2.323 | 1.151 | 4.688 |
| [Composite Housing/Income(0-2) + Education (0-1) Score=2.00] | 0.831 | 0.3569 | 0.131 | 1.531 | 5.421 | 1 | 0.02 | 2.296 | 1.141 | 4.621 |
| [Composite Housing/Income(0-2) + Education (0-1) Score=1.00] | 0.527 | 0.3638 | -0.186 | 1.24 | 2.101 | 1 | 0.147 | 1.694 | 0.831 | 3.457 |
| [Composite Housing/Income(0-2) + Education (0-1) Score=.00] | 0a | . | . | . | . | . | . | 1 | . | . |
| [Confirmed and suspected Covid-19 2 levels=1.00] | -0.003 | 0.1042 | -0.208 | 0.201 | 0.001 | 1 | 0.973 | 0.997 | 0.812 | 1.222 |
| [Confirmed and suspected Covid-19 2 levels=.00] | 0a | . | . | . | . | . | . | 1 | . | . |
| [Limiting_Condition=1.00] | -0.237 | 0.2074 | -0.644 | 0.169 | 1.309 | 1 | 0.252 | 0.789 | 0.525 | 1.184 |
| [Limiting_Condition=.00] | 0a | . | . | . | . | . | . | 1 | . | . |
| [BMI 2 levels including PNTS and DN=2.00] | -0.429 | 0.0928 | -0.611 | -0.248 | 21.404 | 1 | 0 | 0.651 | 0.543 | 0.781 |
| [BMI 2 levels including PNTS and DN=1.00] | 0a | . | . | . | . | . | . | 1 | . | . |
| [Meeting_BEF_categorical=3.00] | 2.106 | 0.1695 | 1.774 | 2.439 | 154.4 | 1 | 0 | 8.218 | 5.895 | 11.457 |
| [Meeting_BEF_categorical=2.00] | 0.451 | 0.1774 | 0.103 | 0.799 | 6.466 | 1 | 0.011 | 1.57 | 1.109 | 2.222 |
| [Meeting_BEF_categorical=1.00] | 1.038 | 0.1895 | 0.666 | 1.409 | 29.965 | 1 | 0 | 2.822 | 1.947 | 4.092 |
| [Meeting_BEF_categorical=.00] | 0a | . | . | . | . | . | . | 1 | . | . |
| [Alcohol_Risk=1.00] | -0.192 | 0.1014 | -0.391 | 0.006 | 3.6 | 1 | 0.058 | 0.825 | 0.676 | 1.006 |
| [Alcohol_Risk=.00] | 0a | . | . | . | . | . | . | 1 | . | . |
| [Smoking baseline binary yes or no=1.00] | -0.301 | 0.1574 | -0.609 | 0.008 | 3.652 | 1 | 0.056 | 0.74 | 0.544 | 1.008 |
| [Smoking baseline binary yes or no=.00] | 0a | . | . | . | . | . | . | 1 | . | . |
| [Time=7] * [Binary Living With Others (Alone=1 With Others=0).=1.00] | 0.322 | 0.2557 | -0.179 | 0.823 | 1.588 | 1 | 0.208 | 1.38 | 0.836 | 2.278 |
| [Time=7] * [Binary Living With Others (Alone=1 With Others=0).=.00] | 0a | . | . | . | . | . | . | 1 | . | . |
| [Time=4] * [Binary Living With Others (Alone=1 With Others=0).=1.00] | 0.746 | 0.2208 | 0.313 | 1.178 | 11.408 | 1 | 0.001 | 2.108 | 1.367 | 3.249 |
| [Time=4] * [Binary Living With Others (Alone=1 With Others=0).=.00] | 0a | . | . | . | . | . | . | 1 | . | . |
| [Time=2] * [Binary Living With Others (Alone=1 With Others=0).=1.00] | 0.26 | 0.1979 | -0.127 | 0.648 | 1.731 | 1 | 0.188 | 1.297 | 0.88 | 1.912 |
| [Time=2] * [Binary Living With Others (Alone=1 With Others=0).=.00] | 0a | . | . | . | . | . | . | 1 | . | . |
| [Time=1] * [Binary Living With Others (Alone=1 With Others=0).=1.00] | 0a | . | . | . | . | . | . | 1 | . | . |
| [Time=1] * [Binary Living With Others (Alone=1 With Others=0).=.00] | 0a | . | . | . | . | . | . | 1 | . | . |
| [Time=7] * [Meeting_BEF_categorical=3.00] | 0.716 | 0.2965 | 0.135 | 1.297 | 5.829 | 1 | 0.016 | 2.046 | 1.144 | 3.658 |
| [Time=7] * [Meeting_BEF_categorical=2.00] | 0.984 | 0.3062 | 0.384 | 1.584 | 10.319 | 1 | 0.001 | 2.675 | 1.467 | 4.874 |
| [Time=7] * [Meeting_BEF_categorical=1.00] | 0.934 | 0.321 | 0.305 | 1.563 | 8.467 | 1 | 0.004 | 2.545 | 1.357 | 4.775 |
| [Time=7] * [Meeting_BEF_categorical=.00] | 0a | . | . | . | . | . | . | 1 | . | . |
| [Time=4] * [Meeting_BEF_categorical=3.00] | 0.361 | 0.2143 | -0.059 | 0.781 | 2.842 | 1 | 0.092 | 1.435 | 0.943 | 2.185 |
| [Time=4] * [Meeting_BEF_categorical=2.00] | 0.424 | 0.2334 | -0.033 | 0.882 | 3.308 | 1 | 0.069 | 1.529 | 0.968 | 2.415 |
| [Time=4] * [Meeting_BEF_categorical=1.00] | 0.469 | 0.2488 | -0.019 | 0.956 | 3.547 | 1 | 0.06 | 1.598 | 0.981 | 2.602 |
| [Time=4] * [Meeting_BEF_categorical=.00] | 0a | . | . | . | . | . | . | 1 | . | . |
| [Time=2] * [Meeting_BEF_categorical=3.00] | 0.107 | 0.1906 | -0.267 | 0.48 | 0.313 | 1 | 0.576 | 1.112 | 0.766 | 1.616 |
| [Time=2] * [Meeting_BEF_categorical=2.00] | 0.416 | 0.1917 | 0.04 | 0.791 | 4.702 | 1 | 0.03 | 1.515 | 1.041 | 2.206 |
| [Time=2] * [Meeting_BEF_categorical=1.00] | 0.072 | 0.2204 | -0.36 | 0.504 | 0.106 | 1 | 0.745 | 1.074 | 0.697 | 1.655 |
| [Time=2] * [Meeting_BEF_categorical=.00] | 0a | . | . | . | . | . | . | 1 | . | . |
| [Time=1] * [Meeting_BEF_categorical=3.00] | 0a | . | . | . | . | . | . | 1 | . | . |
| [Time=1] * [Meeting_BEF_categorical=2.00] | 0a | . | . | . | . | . | . | 1 | . | . |
| [Time=1] * [Meeting_BEF_categorical=1.00] | 0a | . | . | . | . | . | . | 1 | . | . |
| [Time=1] * [Meeting_BEF_categorical=.00] | 0a | . | . | . | . | . | . | 1 | . | . |

**Supplementary table 15 (Complete Case Models –Meeting MVPA Guidelines)**

Tests of Model Effects

| Source | Type III Wald Chi-Square | df | Sig. |
|---|---|---|---|
| (Intercept) | 117.792 | 1 | 0 |
| Time | 36.25 | 3 | 0 |
| gender 2 levels including PNS | 5.615 | 1 | 0.018 |
| Composite_QoL | 34.413 | 1 | 0 |
| age_continuous | 15.048 | 1 | 0 |
| perceived risk at BL including DN | 1.438 | 1 | 0.23 |
| Composite occupation and working from home | 0.27 | 2 | 0.874 |
| Binary Living With Others (Alone=1 With Others=0). | 0.339 | 1 | 0.56 |
| Access to a garden Binary Var | 5.166 | 1 | 0.023 |
| Binary Isolation (Total Isolation=1 All Else = 0). | 33.3 | 1 | 0 |
| ethnicity dichotomised including PNS | 6.353 | 1 | 0.012 |
| Composite Housing/Income(0-2) + Education (0-1) Score | 19.622 | 3 | 0 |
| Confirmed and suspected Covid-19 2 levels | 0.04 | 1 | 0.842 |
| Limiting_Condition | 21.359 | 1 | 0 |
| BMI 2 levels including PNTS and DN | 17.023 | 1 | 0 |
| Meeting_BEF_categorical | 407.286 | 3 | 0 |
| Alcohol_Risk | 3.124 | 1 | 0.077 |
| Smoking baseline binary yes or no | 0.744 | 1 | 0.388 |
| Time * Composite occupation and working from home | 15.151 | 6 | 0.019 |

Dependent Variable: meeting_mvpaSIN

Model: (Intercept), Time, gender 2 levels including PNS, Composite_QoL, age_continuous, perceived risk at BL including DN, Composite occupation and working from home, Binary Living With Others (Alone=1 With Others=0)., Access to a garden Binary Var, Binary Isolation (Total Isolation=1 All Else = 0)., ethnicity dichotomised including PNS, Composite Housing/Income(0-2) + Education (0-1) Score, Confirmed and suspected Covid-19 2 levels, Limiting_Condition, BMI 2 levels including PNTS and DN, Meeting_BEF_categorical, Alcohol_Risk, Smoking baseline binary yes or no, Time * Composite occupation and working from home

| Parameter | B | Std. Error | 95% Wald Confidenc | | Hypothesis Test | | | Exp(B) | 95% Wald Confidenc | |
|---|---|---|---|---|---|---|---|---|---|---|
| | | | Lower | Upper | Wald Chi-! | df | Sig. | | Lower | Upper |
| (Intercept) | -2.631 | 0.3525 | -3.322 | -1.94 | 55.717 | 1 | 0 | 0.072 | 0.036 | 0.144 |
| [Time=7] | -0.092 | 0.116 | -0.32 | 0.135 | 0.632 | 1 | 0.427 | 0.912 | 0.726 | 1.145 |
| [Time=4] | -0.353 | 0.1115 | -0.572 | -0.135 | 10.032 | 1 | 0.002 | 0.702 | 0.564 | 0.874 |
| [Time=2] | -0.135 | 0.1064 | -0.344 | 0.073 | 1.62 | 1 | 0.203 | 0.873 | 0.709 | 1.076 |
| [Time=1] | 0a | . | . | . | . | . | . | 1 | . | . |
| [gender 2 levels including PNS=1.00] | -0.202 | 0.0853 | -0.37 | -0.035 | 5.615 | 1 | 0.018 | 0.817 | 0.691 | 0.966 |
| [gender 2 levels including PNS=.00] | 0a | . | . | . | . | . | . | 1 | . | . |
| Composite_QoL | 0.245 | 0.0418 | 0.163 | 0.327 | 34.413 | 1 | 0 | 1.278 | 1.177 | 1.387 |
| age_continuous | 0.014 | 0.0035 | 0.007 | 0.021 | 15.048 | 1 | 0 | 1.014 | 1.007 | 1.021 |
| [perceived risk at BL including DN=1.00] | -0.076 | 0.0633 | -0.2 | 0.048 | 1.438 | 1 | 0.23 | 0.927 | 0.819 | 1.049 |
| [perceived risk at BL including DN=.00] | 0a | . | . | . | . | . | . | 1 | . | . |
| [Composite occupation and working from home=3.00] | 0.171 | 0.1594 | -0.141 | 0.484 | 1.153 | 1 | 0.283 | 1.187 | 0.868 | 1.622 |
| [Composite occupation and working from home=2.00] | 0.092 | 0.1338 | -0.17 | 0.354 | 0.471 | 1 | 0.492 | 1.096 | 0.843 | 1.425 |
| [Composite occupation and working from home=1.00] | 0a | . | . | . | . | . | . | 1 | . | . |
| [Binary Living With Others (Alone=1 With Others=0).=1.00] | -0.065 | 0.1123 | -0.286 | 0.155 | 0.339 | 1 | 0.56 | 0.937 | 0.752 | 1.167 |
| [Binary Living With Others (Alone=1 With Others=0).=.00] | 0a | . | . | . | . | . | . | 1 | . | . |
| [Access to a garden Binary Var=1.00] | -0.206 | 0.0906 | -0.383 | -0.028 | 5.166 | 1 | 0.023 | 0.814 | 0.682 | 0.972 |
| [Access to a garden Binary Var=.00] | 0a | . | . | . | . | . | . | 1 | . | . |
| [Binary Isolation (Total Isolation=1 All Else = 0).=1.00] | -0.878 | 0.1521 | -1.176 | -0.58 | 33.3 | 1 | 0 | 0.416 | 0.309 | 0.56 |
| [Binary Isolation (Total Isolation=1 All Else = 0).=.00] | 0a | . | . | . | . | . | . | 1 | . | . |
| [ethnicity dichotomised including PNS=1.00] | 0.394 | 0.1563 | 0.088 | 0.7 | 6.353 | 1 | 0.012 | 1.483 | 1.092 | 2.015 |
| [ethnicity dichotomised including PNS=.00] | 0a | . | . | . | . | . | . | 1 | . | . |
| [Composite Housing/Income(0-2) + Education (0-1) Score=3.00] | 0.504 | 0.1916 | 0.128 | 0.879 | 6.912 | 1 | 0.009 | 1.655 | 1.137 | 2.409 |
| [Composite Housing/Income(0-2) + Education (0-1) Score=2.00] | 0.369 | 0.1836 | 0.01 | 0.729 | 4.047 | 1 | 0.044 | 1.447 | 1.01 | 2.073 |
| [Composite Housing/Income(0-2) + Education (0-1) Score=1.00] | 0.061 | 0.1903 | -0.312 | 0.434 | 0.103 | 1 | 0.748 | 1.063 | 0.732 | 1.543 |
| [Composite Housing/Income(0-2) + Education (0-1) Score=.00] | 0a | . | . | . | . | . | . | 1 | . | . |
| [Confirmed and suspected Covid-19 2 levels=1.00] | -0.017 | 0.0834 | -0.18 | 0.147 | 0.04 | 1 | 0.842 | 0.984 | 0.835 | 1.158 |
| [Confirmed and suspected Covid-19 2 levels=.00] | 0a | . | . | . | . | . | . | 1 | . | . |
| [Limiting_Condition=1.00] | -0.629 | 0.1361 | -0.895 | -0.362 | 21.359 | 1 | 0 | 0.533 | 0.408 | 0.696 |
| [Limiting_Condition=.00] | 0a | . | . | . | . | . | . | 1 | . | . |
| [BMI 2 levels including PNTS and DN=2.00] | -0.293 | 0.0709 | -0.431 | -0.154 | 17.023 | 1 | 0 | 0.746 | 0.65 | 0.858 |
| [BMI 2 levels including PNTS and DN=1.00] | 0a | . | . | . | . | . | . | 1 | . | . |
| [Meeting_BEF_categorical=3.00] | 1.552 | 0.1097 | 1.337 | 1.767 | 200.254 | 1 | 0 | 4.722 | 3.809 | 5.855 |
| [Meeting_BEF_categorical=2.00] | 1.662 | 0.0956 | 1.474 | 1.849 | 301.944 | 1 | 0 | 5.269 | 4.368 | 6.355 |
| [Meeting_BEF_categorical=1.00] | 0.239 | 0.1183 | 0.007 | 0.471 | 4.075 | 1 | 0.044 | 1.27 | 1.007 | 1.601 |
| [Meeting_BEF_categorical=.00] | 0a | . | . | . | . | . | . | 1 | . | . |
| [Alcohol_Risk=1.00] | -0.133 | 0.075 | -0.28 | 0.014 | 3.124 | 1 | 0.077 | 0.876 | 0.756 | 1.015 |
| [Alcohol_Risk=.00] | 0a | . | . | . | . | . | . | 1 | . | . |
| [Smoking baseline binary yes or no=1.00] | -0.102 | 0.1178 | -0.332 | 0.129 | 0.744 | 1 | 0.388 | 0.903 | 0.717 | 1.138 |
| [Smoking baseline binary yes or no=.00] | 0a | . | . | . | . | . | . | 1 | . | . |
| [Time=7] * [Composite occupation and working from home=3.00] | -0.429 | 0.1922 | -0.806 | -0.052 | 4.981 | 1 | 0.026 | 0.651 | 0.447 | 0.949 |
| [Time=7] * [Composite occupation and working from home=2.00] | -0.321 | 0.1508 | -0.617 | -0.026 | 4.544 | 1 | 0.033 | 0.725 | 0.54 | 0.974 |
| [Time=7] * [Composite occupation and working from home=1.00] | 0a | . | . | . | . | . | . | 1 | . | . |
| [Time=4] * [Composite occupation and working from home=3.00] | -0.171 | 0.1792 | -0.522 | 0.18 | 0.911 | 1 | 0.34 | 0.843 | 0.593 | 1.197 |
| [Time=4] * [Composite occupation and working from home=2.00] | 0.128 | 0.1386 | -0.144 | 0.399 | 0.848 | 1 | 0.357 | 1.136 | 0.866 | 1.491 |
| [Time=4] * [Composite occupation and working from home=1.00] | 0a | . | . | . | . | . | . | 1 | . | . |
| [Time=2] * [Composite occupation and working from home=3.00] | -0.012 | 0.1671 | -0.34 | 0.315 | 0.005 | 1 | 0.941 | 0.988 | 0.712 | 1.371 |
| [Time=2] * [Composite occupation and working from home=2.00] | 0.041 | 0.1338 | -0.221 | 0.303 | 0.093 | 1 | 0.761 | 1.042 | 0.801 | 1.354 |
| [Time=2] * [Composite occupation and working from home=1.00] | 0a | . | . | . | . | . | . | 1 | . | . |
| [Time=1] * [Composite occupation and working from home=3.00] | 0a | . | . | . | . | . | . | 1 | . | . |
| [Time=1] * [Composite occupation and working from home=2.00] | 0a | . | . | . | . | . | . | 1 | . | . |
| [Time=1] * [Composite occupation and working from home=1.00] | 0a | . | . | . | . | . | . | 1 | . | . |

**Supplementary table 16 (Complete Case Models – Meeting MSA Guidelines)**

Tests of Model Effects

| Source | Type III Wald Chi-Square | df | Sig. |
|---|---|---|---|
| (Intercept) | 17.839 | 1 | 0 |
| Time | 17.19 | 3 | 0.001 |
| gender 2 levels including PNS | 1.238 | 1 | 0.266 |
| Composite_QoL | 30.636 | 1 | 0 |
| age_continuous | 5.296 | 1 | 0.021 |
| perceived risk at BL including DN | 0 | 1 | 0.989 |
| Composite occupation and working from home | 1.728 | 2 | 0.421 |
| Binary Living With Others (Alone=1 With Others=0). | 2.022 | 1 | 0.155 |
| Access to a garden Binary Var | 0.215 | 1 | 0.643 |
| Binary Isolation (Total Isolation=1 All Else = 0). | 0.024 | 1 | 0.878 |
| ethnicity dichotomised including PNS | 4.409 | 1 | 0.036 |
| Composite Housing/Income(0-2) + Education (0-1) Score | 1.936 | 3 | 0.586 |
| Confirmed and suspected Covid-19 2 levels | 0.069 | 1 | 0.792 |
| Limiting_Condition | 0.737 | 1 | 0.39 |
| BMI 2 levels including PNTS and DN | 23.795 | 1 | 0 |
| Meeting_BEF_categorical | 513.88 | 3 | 0 |
| Alcohol_Risk | 9.671 | 1 | 0.002 |
| Smoking baseline binary yes or no | 4.264 | 1 | 0.039 |
| Time * gender 2 levels including PNS | 10.02 | 3 | 0.018 |
| Time * age_continuous | 10.882 | 3 | 0.012 |

Dependent Variable: meeting_msaSIN

Model: (Intercept), Time, gender 2 levels including PNS, Composite_QoL, age_continuous, perceived risk at BL including DN, Composite occupation and working from home, Binary Living With Others (Alone=1 With Others=0)., Access to a garden Binary Var, Binary Isolation (Total Isolation=1 All Else = 0)., ethnicity dichotomised including PNS, Composite Housing/Income(0-2) + Education (0-1) Score, Confirmed and suspected Covid-19 2 levels, Limiting_Condition, BMI 2 levels including PNTS and DN, Meeting_BEF_categorical, Alcohol_Risk, Smoking baseline binary yes or no, Time * gender 2 levels including PNS, Time * age_continuous

| Parameter | B | Std. Error | 95% Wald Confidenc | | Hypothesis Test | | | Exp(B) | 95% Wald Confidenc | |
|---|---|---|---|---|---|---|---|---|---|---|
| | | | Lower | Upper | Wald Chi-! | df | Sig. | | Lower | Upper |
| (Intercept) | -1.097 | 0.4683 | -2.015 | -0.179 | 5.489 | 1 | 0.019 | 0.334 | 0.133 | 0.836 |
| [Time=7] | -0.838 | 0.2807 | -1.389 | -0.288 | 8.92 | 1 | 0.003 | 0.432 | 0.249 | 0.75 |
| [Time=4] | -0.498 | 0.251 | -0.99 | -0.006 | 3.941 | 1 | 0.047 | 0.607 | 0.371 | 0.994 |
| [Time=2] | -0.21 | 0.2243 | -0.65 | 0.229 | 0.878 | 1 | 0.349 | 0.81 | 0.522 | 1.258 |
| [Time=1] | 0a | . | . | . | . | . | . | 1 | . | . |
| [gender 2 levels including PNS=1.00] | 0.123 | 0.124 | -0.12 | 0.366 | 0.986 | 1 | 0.321 | 1.131 | 0.887 | 1.442 |
| [gender 2 levels including PNS=.00] | 0a | . | . | . | . | . | . | 1 | . | . |
| Composite_QoL | 0.251 | 0.0454 | 0.162 | 0.34 | 30.636 | 1 | 0 | 1.286 | 1.176 | 1.405 |
| age_continuous | -0.016 | 0.0044 | -0.025 | -0.008 | 13.855 | 1 | 0 | 0.984 | 0.975 | 0.992 |
| [perceived risk at BL including DN=1.00] | -0.001 | 0.0655 | -0.129 | 0.127 | 0 | 1 | 0.989 | 0.999 | 0.879 | 1.136 |
| [perceived risk at BL including DN=.00] | 0a | . | . | . | . | . | . | 1 | . | . |
| [Composite occupation and working from home=3.00] | -0.149 | 0.1386 | -0.42 | 0.123 | 1.149 | 1 | 0.284 | 0.862 | 0.657 | 1.131 |
| [Composite occupation and working from home=2.00] | 0 | 0.1195 | -0.234 | 0.234 | 0 | 1 | 0.999 | 1 | 0.791 | 1.264 |
| [Composite occupation and working from home=1.00] | 0a | . | . | . | . | . | . | 1 | . | . |
| [Binary Living With Others (Alone=1 With Others=0).=1.00] | -0.177 | 0.1247 | -0.422 | 0.067 | 2.022 | 1 | 0.155 | 0.838 | 0.656 | 1.069 |
| [Binary Living With Others (Alone=1 With Others=0).=.00] | 0a | . | . | . | . | . | . | 1 | . | . |
| [Access to a garden Binary Var=1.00] | -0.047 | 0.1011 | -0.245 | 0.151 | 0.215 | 1 | 0.643 | 0.954 | 0.783 | 1.163 |
| [Access to a garden Binary Var=.00] | 0a | . | . | . | . | . | . | 1 | . | . |
| [Binary Isolation (Total Isolation=1 All Else = 0).=1.00] | -0.023 | 0.1478 | -0.312 | 0.267 | 0.024 | 1 | 0.878 | 0.978 | 0.732 | 1.306 |
| [Binary Isolation (Total Isolation=1 All Else = 0).=.00] | 0a | . | . | . | . | . | . | 1 | . | . |
| [ethnicity dichotomised including PNS=1.00] | -0.414 | 0.1971 | -0.8 | -0.028 | 4.409 | 1 | 0.036 | 0.661 | 0.449 | 0.973 |
| [ethnicity dichotomised including PNS=.00] | 0a | . | . | . | . | . | . | 1 | . | . |
| [Composite Housing/Income(0-2) + Education (0-1) Score=3.00] | 0.365 | 0.3182 | -0.259 | 0.989 | 1.315 | 1 | 0.251 | 1.44 | 0.772 | 2.687 |
| [Composite Housing/Income(0-2) + Education (0-1) Score=2.00] | 0.331 | 0.3143 | -0.285 | 0.947 | 1.109 | 1 | 0.292 | 1.392 | 0.752 | 2.578 |
| [Composite Housing/Income(0-2) + Education (0-1) Score=1.00] | 0.251 | 0.3187 | -0.374 | 0.875 | 0.618 | 1 | 0.432 | 1.285 | 0.688 | 2.399 |
| [Composite Housing/Income(0-2) + Education (0-1) Score=.00] | 0a | . | . | . | . | . | . | 1 | . | . |
| [Confirmed and suspected Covid-19 2 levels=1.00] | -0.023 | 0.0872 | -0.194 | 0.148 | 0.069 | 1 | 0.792 | 0.977 | 0.824 | 1.159 |
| [Confirmed and suspected Covid-19 2 levels=.00] | 0a | . | . | . | . | . | . | 1 | . | . |
| [Limiting_Condition=1.00] | -0.137 | 0.1598 | -0.45 | 0.176 | 0.737 | 1 | 0.39 | 0.872 | 0.637 | 1.192 |
| [Limiting_Condition=.00] | 0a | . | . | . | . | . | . | 1 | . | . |
| [BMI 2 levels including PNTS and DN=2.00] | -0.375 | 0.0768 | -0.525 | -0.224 | 23.795 | 1 | 0 | 0.688 | 0.591 | 0.799 |
| [BMI 2 levels including PNTS and DN=1.00] | 0a | . | . | . | . | . | . | 1 | . | . |
| [Meeting_BEF_categorical=3.00] | 2.056 | 0.1171 | 1.827 | 2.286 | 308.428 | 1 | 0 | 7.817 | 6.214 | 9.833 |
| [Meeting_BEF_categorical=2.00] | 0.078 | 0.1157 | -0.149 | 0.305 | 0.453 | 1 | 0.501 | 1.081 | 0.862 | 1.356 |
| [Meeting_BEF_categorical=1.00] | 1.984 | 0.1226 | 1.744 | 2.225 | 261.949 | 1 | 0 | 7.274 | 5.72 | 9.25 |
| [Meeting_BEF_categorical=.00] | 0a | . | . | . | . | . | . | 1 | . | . |
| [Alcohol_Risk=1.00] | -0.241 | 0.0773 | -0.392 | -0.089 | 9.671 | 1 | 0.002 | 0.786 | 0.676 | 0.915 |
| [Alcohol_Risk=.00] | 0a | . | . | . | . | . | . | 1 | . | . |
| [Smoking baseline binary yes or no=1.00] | -0.264 | 0.1281 | -0.516 | -0.013 | 4.264 | 1 | 0.039 | 0.768 | 0.597 | 0.987 |
| [Smoking baseline binary yes or no=.00] | 0a | . | . | . | . | . | . | 1 | . | . |
| [Time=7] * [gender 2 levels including PNS=1.00] | -0.326 | 0.1477 | -0.616 | -0.037 | 4.877 | 1 | 0.027 | 0.722 | 0.54 | 0.964 |
| [Time=7] * [gender 2 levels including PNS=.00] | 0a | . | . | . | . | . | . | 1 | . | . |
| [Time=4] * [gender 2 levels including PNS=1.00] | -0.424 | 0.1374 | -0.693 | -0.154 | 9.507 | 1 | 0.002 | 0.655 | 0.5 | 0.857 |
| [Time=4] * [gender 2 levels including PNS=.00] | 0a | . | . | . | . | . | . | 1 | . | . |
| [Time=2] * [gender 2 levels including PNS=1.00] | -0.173 | 0.1239 | -0.416 | 0.07 | 1.956 | 1 | 0.162 | 0.841 | 0.66 | 1.072 |
| [Time=2] * [gender 2 levels including PNS=.00] | 0a | . | . | . | . | . | . | 1 | . | . |
| [Time=1] * [gender 2 levels including PNS=1.00] | 0a | . | . | . | . | . | . | 1 | . | . |
| [Time=1] * [gender 2 levels including PNS=.00] | 0a | . | . | . | . | . | . | 1 | . | . |
| [Time=7] * age_continuous | 0.015 | 0.0047 | 0.006 | 0.024 | 9.788 | 1 | 0.002 | 1.015 | 1.006 | 1.024 |
| [Time=4] * age_continuous | 0.011 | 0.0043 | 0.002 | 0.019 | 6.206 | 1 | 0.013 | 1.011 | 1.002 | 1.02 |
| [Time=2] * age_continuous | 0.006 | 0.004 | -0.002 | 0.013 | 2.018 | 1 | 0.155 | 1.006 | 0.998 | 1.013 |
| [Time=1] * age_continuous | 0a | . | . | . | . | . | . | 1 | . | . |
| (Scale) | 1 | | | | | | | | | |

**Supplementary table 17 (Count of PA sessions Dependent Variable Pairwise Comparisons)**

| Estimates | | | | |
|---|---|---|---|---|
| Time | Mean | Std. Error | 95% Wald Confidenc | |
| | | | Lower | Upper |
| 7 | 4.7352 | 0.09261 | 4.5571 | 4.9202 |
| 4 | 4.8115 | 0.08759 | 4.6429 | 4.9863 |
| 2 | 5.1067 | 0.08946 | 4.9343 | 5.2851 |
| 1 | 4.8911 | 0.08285 | 4.7314 | 5.0562 |

| Pairwise Comparisons | | | | | | | |
|---|---|---|---|---|---|---|---|
| (I) Time | (J) Time | Mean Diff | Std. Error | df | Sequentia | 95% Wald Confidenc | |
| | | | | | | Lower | Upper |
| 7 | 4 | -0.0763 | 0.07907 | 1 | 0.527 | -0.251 | 0.0983 |
| | 2 | -.3715b | 0.08379 | 1 | 0 | -0.592 | -0.1511 |
| | 1 | -0.156 | 0.08862 | 1 | 0.217 | -0.3676 | 0.0556 |
| 4 | 7 | 0.0763 | 0.07907 | 1 | 0.527 | -0.0983 | 0.251 |
| | 2 | -.2952b | 0.07581 | 1 | 0 | -0.4899 | -0.1005 |
| | 1 | -0.0796 | 0.0788 | 1 | 0.527 | -0.2559 | 0.0966 |
| 2 | 7 | .3715b | 0.08379 | 1 | 0 | 0.1511 | 0.592 |
| | 4 | .2952b | 0.07581 | 1 | 0 | 0.1005 | 0.4899 |
| | 1 | .2156b | 0.07037 | 1 | 0.009 | 0.0403 | 0.3909 |
| 1 | 7 | 0.156 | 0.08862 | 1 | 0.217 | -0.0556 | 0.3676 |
| | 4 | 0.0796 | 0.0788 | 1 | 0.527 | -0.0966 | 0.2559 |
| | 2 | -.2156b | 0.07037 | 1 | 0.009 | -0.3909 | -0.0403 |
| Pairwise comparisons of estimated marginal means based on the original scale of de | | | | | | | |
| a Confidence interval bounds are approximate. | | | | | | | |
| b The mean difference is significant at the .05 level. | | | | | | | |

**Supplementary table 18 (Count of PA sessions – Including Pre-Covid PA*Time Interaction Term)**

Tests of Model Effects

| Source | Type III | | |
|---|---|---|---|
| | Wald Chi-Square | df | Sig. |
| (Intercept) | 126.645 | 1 | 0 |
| Time | 24.524 | 3 | 0 |
| gender 2 levels including PNS | 0.122 | 1 | 0.727 |
| Composite_QoL | 51.726 | 1 | 0 |
| age_continuous | 1.939 | 1 | 0.164 |
| Composite occupation and working from home | 2.828 | 2 | 0.243 |
| Binary Living With Others (Alone=1 With Others=0). | 0.233 | 1 | 0.629 |
| Access to a garden Binary Var | 0.158 | 1 | 0.691 |
| Binary Isolation (Total Isolation=1 All Else = 0). | 33.597 | 1 | 0 |
| ethnicity dichotomised including PNS | 0.239 | 1 | 0.625 |
| Composite Housing/Income(0-2) + Education (0-1) Score | 10.067 | 3 | 0.018 |
| Confirmed and suspected Covid-19 2 levels | 0.068 | 1 | 0.795 |
| Limiting_Condition | 11.248 | 1 | 0.001 |
| BMI 2 levels including PNTS and DN | 29.507 | 1 | 0 |
| Time * Meeting_BEF_categorical | 440.576 | 12 | 0 |

Dependent Variable: PA_Count

Model: (Intercept), Time, gender 2 levels including PNS, Composite_QoL, age_continuous, Composite occupation and working from home, Binary Living With Others (Alone=1 With Others=0)., Access to a garden Binary Var, Binary Isolation (Total Isolation=1 All Else = 0)., ethnicity dichotomised including PNS, Composite Housing/Income(0-2) + Education (0-1) Score, Confirmed and suspected Covid-19 2 levels, Limiting Condition, BMI 2 levels including PNTS and DN, Time * Meeting_BEF_categorical

| Parameter | B | Std. Error | 95% Wald Confidenc | | Hypothesis Test | | | Exp(B) | 95% Wald Confidenc | |
|---|---|---|---|---|---|---|---|---|---|---|
| | | | Lower | Upper | Wald Chi- | df | Sig. | | Lower | Upper |
| (Intercept) | 0.812 | 0.1312 | 0.555 | 1.069 | 38.266 | 1 | 0 | 2.252 | 1.741 | 2.912 |
| [Time=7] | -0.119 | 0.0405 | -0.198 | -0.04 | 8.636 | 1 | 0.003 | 0.888 | 0.82 | 0.961 |
| [Time=4] | -0.099 | 0.0334 | -0.164 | -0.033 | 8.745 | 1 | 0.003 | 0.906 | 0.849 | 0.967 |
| [Time=2] | 0.067 | 0.0286 | 0.01 | 0.123 | 5.397 | 1 | 0.02 | 1.069 | 1.01 | 1.13 |
| [Time=1] | 0a | . | . | . | . | . | . | 1 | . | . |
| [gender 2 levels including PNS=1.00] | -0.01 | 0.0282 | -0.065 | 0.045 | 0.122 | 1 | 0.727 | 0.99 | 0.937 | 1.046 |
| [gender 2 levels including PNS=.00] | 0a | . | . | . | . | . | . | 1 | . | . |
| Composite_QoL | 0.087 | 0.0121 | 0.064 | 0.111 | 51.726 | 1 | 0 | 1.091 | 1.066 | 1.118 |
| age_continuous | 0.002 | 0.0012 | -0.001 | 0.004 | 1.939 | 1 | 0.164 | 1.002 | 0.999 | 1.004 |
| [Composite occupation and working from home=3.00] | -0.052 | 0.0428 | -0.136 | 0.032 | 1.471 | 1 | 0.225 | 0.949 | 0.873 | 1.033 |
| [Composite occupation and working from home=2.00] | 0.01 | 0.0359 | -0.06 | 0.081 | 0.085 | 1 | 0.77 | 1.011 | 0.942 | 1.084 |
| [Composite occupation and working from home=1.00] | 0a | . | . | . | . | . | . | 1 | . | . |
| [Binary Living With Others (Alone=1 With Others=0).=1.00] | -0.019 | 0.0384 | -0.094 | 0.057 | 0.233 | 1 | 0.629 | 0.982 | 0.91 | 1.058 |
| [Binary Living With Others (Alone=1 With Others=0).=.00] | 0a | . | . | . | . | . | . | 1 | . | . |
| [Access to a garden Binary Var=1.00] | 0.012 | 0.0303 | -0.047 | 0.072 | 0.158 | 1 | 0.691 | 1.012 | 0.954 | 1.074 |
| [Access to a garden Binary Var=.00] | 0a | . | . | . | . | . | . | 1 | . | . |
| [Binary Isolation (Total Isolation=1 All Else = 0).=1.00] | -0.307 | 0.053 | -0.411 | -0.203 | 33.597 | 1 | 0 | 0.736 | 0.663 | 0.816 |
| [Binary Isolation (Total Isolation=1 All Else = 0).=.00] | 0a | . | . | . | . | . | . | 1 | . | . |
| [ethnicity dichotomised including PNS=1.00] | 0.027 | 0.0544 | -0.08 | 0.133 | 0.239 | 1 | 0.625 | 1.027 | 0.923 | 1.142 |
| [ethnicity dichotomised including PNS=.00] | 0a | . | . | . | . | . | . | 1 | . | . |
| [Composite Housing/Income(0-2) + Education (0-1) Score=3.00] | 0.149 | 0.0873 | -0.022 | 0.32 | 2.924 | 1 | 0.087 | 1.161 | 0.978 | 1.378 |
| [Composite Housing/Income(0-2) + Education (0-1) Score=2.00] | 0.147 | 0.0873 | -0.024 | 0.318 | 2.841 | 1 | 0.092 | 1.159 | 0.976 | 1.375 |
| [Composite Housing/Income(0-2) + Education (0-1) Score=1.00] | 0.054 | 0.0894 | -0.121 | 0.229 | 0.367 | 1 | 0.545 | 1.056 | 0.886 | 1.258 |
| [Composite Housing/Income(0-2) + Education (0-1) Score=.00] | 0a | . | . | . | . | . | . | 1 | . | . |
| [Confirmed and suspected Covid-19 2 levels=1.00] | -0.006 | 0.0246 | -0.055 | 0.042 | 0.068 | 1 | 0.795 | 0.994 | 0.947 | 1.043 |
| [Confirmed and suspected Covid-19 2 levels=.00] | 0a | . | . | . | . | . | . | 1 | . | . |
| [Limiting_Condition=1.00] | -0.189 | 0.0564 | -0.3 | -0.079 | 11.248 | 1 | 0.001 | 0.828 | 0.741 | 0.924 |
| [Limiting_Condition=.00] | 0a | . | . | . | . | . | . | 1 | . | . |
| [BMI 2 levels including PNTS and DN=2.00] | -0.115 | 0.0212 | -0.157 | -0.074 | 29.507 | 1 | 0 | 0.891 | 0.855 | 0.929 |
| [BMI 2 levels including PNTS and DN=1.00] | 0a | . | . | . | . | . | . | 1 | . | . |
| [Time=7] * [Meeting_BEF_categorical=3.00] | 0.8 | 0.0499 | 0.702 | 0.898 | 256.988 | 1 | 0 | 2.225 | 2.018 | 2.454 |
| [Time=7] * [Meeting_BEF_categorical=2.00] | 0.582 | 0.0487 | 0.487 | 0.678 | 143.173 | 1 | 0 | 1.79 | 1.627 | 1.969 |
| [Time=7] * [Meeting_BEF_categorical=1.00] | 0.494 | 0.056 | 0.384 | 0.604 | 77.778 | 1 | 0 | 1.639 | 1.469 | 1.83 |
| [Time=7] * [Meeting_BEF_categorical=.00] | 0a | . | . | . | . | . | . | 1 | . | . |
| [Time=4] * [Meeting_BEF_categorical=3.00] | 0.754 | 0.047 | 0.662 | 0.846 | 257.786 | 1 | 0 | 2.125 | 1.939 | 2.33 |
| [Time=4] * [Meeting_BEF_categorical=2.00] | 0.565 | 0.0455 | 0.475 | 0.654 | 153.83 | 1 | 0 | 1.759 | 1.609 | 1.923 |
| [Time=4] * [Meeting_BEF_categorical=1.00] | 0.489 | 0.0511 | 0.389 | 0.589 | 91.666 | 1 | 0 | 1.631 | 1.475 | 1.802 |
| [Time=4] * [Meeting_BEF_categorical=.00] | 0a | . | . | . | . | . | . | 1 | . | . |
| [Time=2] * [Meeting_BEF_categorical=3.00] | 0.612 | 0.0455 | 0.523 | 0.702 | 181.593 | 1 | 0 | 1.845 | 1.688 | 2.017 |
| [Time=2] * [Meeting_BEF_categorical=2.00] | 0.473 | 0.0433 | 0.388 | 0.558 | 119.05 | 1 | 0 | 1.604 | 1.474 | 1.747 |
| [Time=2] * [Meeting_BEF_categorical=1.00] | 0.334 | 0.0489 | 0.238 | 0.43 | 46.721 | 1 | 0 | 1.397 | 1.269 | 1.537 |
| [Time=2] * [Meeting_BEF_categorical=.00] | 0a | . | . | . | . | . | . | 1 | . | . |
| [Time=1] * [Meeting_BEF_categorical=3.00] | 0.711 | 0.0427 | 0.627 | 0.795 | 276.861 | 1 | 0 | 2.036 | 1.872 | 2.214 |
| [Time=1] * [Meeting_BEF_categorical=2.00] | 0.476 | 0.0409 | 0.396 | 0.557 | 135.677 | 1 | 0 | 1.61 | 1.486 | 1.745 |
| [Time=1] * [Meeting_BEF_categorical=1.00] | 0.433 | 0.046 | 0.343 | 0.523 | 88.517 | 1 | 0 | 1.542 | 1.409 | 1.688 |
| [Time=1] * [Meeting_BEF_categorical=.00] | 0a | . | . | . | . | . | . | 1 | . | . |

**Supplementary table 19 (Count of PA sessions – Age*Time Interaction Term)**

Tests of Model Effects

| Source | Type III Wald Chi-Square | df | Sig. |
|---|---|---|---|
| (Intercept) | 91.521 | 1 | 0 |
| Time | 10.939 | 3 | 0.012 |
| gender 2 levels including PNS | 0.374 | 1 | 0.541 |
| Composite_QoL | 47.166 | 1 | 0 |
| age_continuous | 1.643 | 1 | 0.2 |
| Composite occupation and working from home | 3.992 | 2 | 0.136 |
| Binary Living With Others (Alone=1 With Others=0). | 0.048 | 1 | 0.827 |
| Access to a garden Binary Var | 0.19 | 1 | 0.663 |
| Binary Isolation (Total Isolation=1 All Else = 0). | 31.778 | 1 | 0 |
| ethnicity dichotomised including PNS | 0.739 | 1 | 0.39 |
| Composite Housing/Income(0-2) + Education (0-1) Score | 17.766 | 3 | 0 |
| Confirmed and suspected Covid-19 2 levels | 0.026 | 1 | 0.872 |
| Limiting_Condition | 23.043 | 1 | 0 |
| BMI 2 levels including PNTS and DN | 54.83 | 1 | 0 |
| Time * age_continuous | 8.821 | 3 | 0.032 |

Dependent Variable: PA_Count

Model: (Intercept), Time, gender 2 levels including PNS, Composite_QoL, age_continuous, Composite occupation and working from home, Binary Living With Others (Alone=1 With Others=0)., Access to a garden Binary Var, Binary Isolation (Total Isolation=1 All Else = 0)., ethnicity dichotomised including PNS, Composite Housing/Income(0-2) + Education (0-1) Score, Confirmed and suspected Covid-19 2 levels, Limiting_Condition, BMI 2 levels including PNTS and DN, Time * age_continuous

| Parameter | B | Std. Error | 95% Wald Confidenc | | Hypothesis Test | | | Exp(B) | 95% Wald Confidenc | |
|---|---|---|---|---|---|---|---|---|---|---|
| | | | Lower | Upper | Wald Chi-! | df | Sig. | | Lower | Upper |
| (Intercept) | 1.266 | 0.1427 | 0.986 | 1.546 | 78.664 | 1 | 0 | 3.546 | 2.681 | 4.691 |
| [Time=7] | -0.204 | 0.0714 | -0.344 | -0.064 | 8.19 | 1 | 0.004 | 0.815 | 0.709 | 0.938 |
| [Time=4] | -0.026 | 0.0595 | -0.143 | 0.09 | 0.194 | 1 | 0.66 | 0.974 | 0.867 | 1.095 |
| [Time=2] | -0.001 | 0.052 | -0.103 | 0.101 | 0 | 1 | 0.986 | 0.999 | 0.902 | 1.106 |
| [Time=1] | 0a | . | . | . | . | . | . | 1 | . | . |
| [gender 2 levels including PNS=1.00] | -0.019 | 0.031 | -0.08 | 0.042 | 0.374 | 1 | 0.541 | 0.981 | 0.923 | 1.043 |
| [gender 2 levels including PNS=.00] | 0a | . | . | . | . | . | . | 1 | . | . |
| Composite_QoL | 0.088 | 0.0129 | 0.063 | 0.114 | 47.166 | 1 | 0 | 1.092 | 1.065 | 1.12 |
| age_continuous | 0.001 | 0.0013 | -0.002 | 0.003 | 0.483 | 1 | 0.487 | 1.001 | 0.998 | 1.003 |
| [Composite occupation and working from home=3.00] | -0.086 | 0.0488 | -0.182 | 0.01 | 3.105 | 1 | 0.078 | 0.918 | 0.834 | 1.01 |
| [Composite occupation and working from home=2.00] | -0.009 | 0.0404 | -0.088 | 0.07 | 0.05 | 1 | 0.823 | 0.991 | 0.915 | 1.073 |
| [Composite occupation and working from home=1.00] | 0a | . | . | . | . | . | . | 1 | . | . |
| [Binary Living With Others (Alone=1 With Others=0).=1.00] | -0.009 | 0.0433 | -0.094 | 0.075 | 0.048 | 1 | 0.827 | 0.991 | 0.91 | 1.078 |
| [Binary Living With Others (Alone=1 With Others=0).=.00] | 0a | . | . | . | . | . | . | 1 | . | . |
| [Access to a garden Binary Var=1.00] | 0.015 | 0.0334 | -0.051 | 0.08 | 0.19 | 1 | 0.663 | 1.015 | 0.95 | 1.083 |
| [Access to a garden Binary Var=.00] | 0a | . | . | . | . | . | . | 1 | . | . |
| [Binary Isolation (Total Isolation=1 All Else = 0).=1.00] | -0.306 | 0.0543 | -0.413 | -0.2 | 31.778 | 1 | 0 | 0.736 | 0.662 | 0.819 |
| [Binary Isolation (Total Isolation=1 All Else = 0).=.00] | 0a | . | . | . | . | . | . | 1 | . | . |
| [ethnicity dichotomised including PNS=1.00] | -0.053 | 0.0617 | -0.174 | 0.068 | 0.739 | 1 | 0.39 | 0.948 | 0.84 | 1.07 |
| [ethnicity dichotomised including PNS=.00] | 0a | . | . | . | . | . | . | 1 | . | . |
| [Composite Housing/Income(0-2) + Education (0-1) Score=3.00] | 0.232 | 0.0985 | 0.039 | 0.425 | 5.56 | 1 | 0.018 | 1.261 | 1.04 | 1.53 |
| [Composite Housing/Income(0-2) + Education (0-1) Score=2.00] | 0.19 | 0.0979 | -0.002 | 0.382 | 3.769 | 1 | 0.052 | 1.209 | 0.998 | 1.465 |
| [Composite Housing/Income(0-2) + Education (0-1) Score=1.00] | 0.076 | 0.1002 | -0.12 | 0.272 | 0.574 | 1 | 0.449 | 1.079 | 0.887 | 1.313 |
| [Composite Housing/Income(0-2) + Education (0-1) Score=.00] | 0a | . | . | . | . | . | . | 1 | . | . |
| [Confirmed and suspected Covid-19 2 levels=1.00] | -0.004 | 0.0258 | -0.055 | 0.046 | 0.026 | 1 | 0.872 | 0.996 | 0.947 | 1.047 |
| [Confirmed and suspected Covid-19 2 levels=.00] | 0a | . | . | . | . | . | . | 1 | . | . |
| [Limiting_Condition=1.00] | -0.306 | 0.0637 | -0.431 | -0.181 | 23.043 | 1 | 0 | 0.737 | 0.65 | 0.834 |
| [Limiting_Condition=.00] | 0a | . | . | . | . | . | . | 1 | . | . |
| [BMI 2 levels including PNTS and DN=2.00] | -0.164 | 0.0222 | -0.208 | -0.121 | 54.83 | 1 | 0 | 0.848 | 0.812 | 0.886 |
| [BMI 2 levels including PNTS and DN=1.00] | 0a | . | . | . | . | . | . | 1 | . | . |
| [Time=7] * age_continuous | 0.003 | 0.0013 | 0 | 0.005 | 4.779 | 1 | 0.029 | 1.003 | 1 | 1.005 |
| [Time=4] * age_continuous | -0.001 | 0.0011 | -0.003 | 0.002 | 0.271 | 1 | 0.603 | 0.999 | 0.997 | 1.002 |
| [Time=2] * age_continuous | 0 | 0.001 | -0.001 | 0.002 | 0.24 | 1 | 0.624 | 1 | 0.999 | 1.002 |
| [Time=1] * age_continuous | 0a | . | . | . | . | . | . | 1 | . | . |

**Supplementary table 20 (Count of PA sessions – Gender*Time Interaction Term)**

Tests of Model Effects

| Source | Type III Wald Chi-Square | df | Sig. |
|---|---|---|---|
| (Intercept) | 90.374 | 1 | 0 |
| Time | 19.763 | 3 | 0 |
| gender 2 levels including PNS | 0.838 | 1 | 0.36 |
| Composite_QoL | 49.947 | 1 | 0 |
| age_continuous | 1.823 | 1 | 0.177 |
| Composite occupation and working from home | 3.908 | 2 | 0.142 |
| Binary Living With Others (Alone=1 With Others=0). | 0.042 | 1 | 0.838 |
| Access to a garden Binary Var | 0.203 | 1 | 0.652 |
| Binary Isolation (Total Isolation=1 All Else = 0). | 32.083 | 1 | 0 |
| ethnicity dichotomised including PNS | 0.818 | 1 | 0.366 |
| Composite Housing/Income(0-2) + Education (0-1) Score | 17.642 | 3 | 0.001 |
| Confirmed and suspected Covid-19 2 levels | 0.06 | 1 | 0.807 |
| Limiting_Condition | 22.928 | 1 | 0 |
| BMI 2 levels including PNTS and DN | 55.536 | 1 | 0 |
| Time * gender 2 levels including PNS | 8.871 | 3 | 0.031 |

Dependent Variable: PA_Count
Model: (Intercept), Time, gender 2 levels including PNS, Composite_QoL, age_continuous, Composite occupation and working from home, Binary Living With Others (Alone=1 With Others=0)., Access to a garden Binary Var, Binary Isolation (Total Isolation=1 All Else = 0)., ethnicity dichotomised including PNS, Composite Housing/Income(0-2) + Education (0-1) Score, Confirmed and suspected Covid-19 2 levels, Limiting_Condition, BMI 2 levels including PNTS and DN, Time * gender 2 levels including PNS

| Parameter | B | Std. Error | 95% Wald Confidenc |  | Hypothesis Test |  |  | Exp(B) | 95% Wald Confidenc |  |
|---|---|---|---|---|---|---|---|---|---|---|
|  |  |  | Lower | Upper | Wald Chi- | df | Sig. |  | Lower | Upper |
| (Intercept) | 1.192 | 0.1417 | 0.915 | 1.47 | 70.806 | 1 | 0 | 3.295 | 2.496 | 4.35 |
| [Time=7] | -0.012 | 0.0342 | -0.079 | 0.055 | 0.117 | 1 | 0.733 | 0.988 | 0.924 | 1.057 |
| [Time=4] | 0.017 | 0.0311 | -0.044 | 0.078 | 0.307 | 1 | 0.579 | 1.017 | 0.957 | 1.081 |
| [Time=2] | 0.043 | 0.0262 | -0.008 | 0.095 | 2.711 | 1 | 0.1 | 1.044 | 0.992 | 1.099 |
| [Time=1] | 0a | . | . | . | . | . | . | 1 | . | . |
| [gender 2 levels including PNS=1.00] | 0.021 | 0.0356 | -0.049 | 0.09 | 0.333 | 1 | 0.564 | 1.021 | 0.952 | 1.095 |
| [gender 2 levels including PNS=.00] | 0a | . | . | . | . | . | . | 1 | . | . |
| Composite_QoL | 0.091 | 0.0129 | 0.066 | 0.116 | 49.947 | 1 | 0 | 1.095 | 1.068 | 1.123 |
| age_continuous | 0.002 | 0.0012 | -0.001 | 0.004 | 1.823 | 1 | 0.177 | 1.002 | 0.999 | 1.004 |
| [Composite occupation and working from home=3.00] | -0.085 | 0.0487 | -0.181 | 0.01 | 3.061 | 1 | 0.08 | 0.918 | 0.835 | 1.01 |
| [Composite occupation and working from home=2.00] | -0.009 | 0.0404 | -0.088 | 0.07 | 0.053 | 1 | 0.818 | 0.991 | 0.915 | 1.072 |
| [Composite occupation and working from home=1.00] | 0a | . | . | . | . | . | . | 1 | . | . |
| [Binary Living With Others (Alone=1 With Others=0).=1.00] | -0.009 | 0.0432 | -0.094 | 0.076 | 0.042 | 1 | 0.838 | 0.991 | 0.911 | 1.079 |
| [Binary Living With Others (Alone=1 With Others=0).=.00] | 0a | . | . | . | . | . | . | 1 | . | . |
| [Access to a garden Binary Var=1.00] | 0.015 | 0.0334 | -0.05 | 0.08 | 0.203 | 1 | 0.652 | 1.015 | 0.951 | 1.084 |
| [Access to a garden Binary Var=.00] | 0a | . | . | . | . | . | . | 1 | . | . |
| [Binary Isolation (Total Isolation=1 All Else = 0).=1.00] | -0.309 | 0.0545 | -0.415 | -0.202 | 32.083 | 1 | 0 | 0.734 | 0.66 | 0.817 |
| [Binary Isolation (Total Isolation=1 All Else = 0).=.00] | 0a | . | . | . | . | . | . | 1 | . | . |
| [ethnicity dichotomised including PNS=1.00] | -0.056 | 0.0615 | -0.176 | 0.065 | 0.818 | 1 | 0.366 | 0.946 | 0.839 | 1.067 |
| [ethnicity dichotomised including PNS=.00] | 0a | . | . | . | . | . | . | 1 | . | . |
| [Composite Housing/Income(0-2) + Education (0-1) Score=3.00] | 0.233 | 0.0985 | 0.04 | 0.426 | 5.583 | 1 | 0.018 | 1.262 | 1.04 | 1.531 |
| [Composite Housing/Income(0-2) + Education (0-1) Score=2.00] | 0.193 | 0.0978 | 0.001 | 0.385 | 3.884 | 1 | 0.049 | 1.213 | 1.001 | 1.469 |
| [Composite Housing/Income(0-2) + Education (0-1) Score=1.00] | 0.078 | 0.1002 | -0.119 | 0.274 | 0.604 | 1 | 0.437 | 1.081 | 0.888 | 1.316 |
| [Composite Housing/Income(0-2) + Education (0-1) Score=.00] | 0a | . | . | . | . | . | . | 1 | . | . |
| [Confirmed and suspected Covid-19 2 levels=1.00] | -0.006 | 0.0258 | -0.057 | 0.044 | 0.06 | 1 | 0.807 | 0.994 | 0.945 | 1.045 |
| [Confirmed and suspected Covid-19 2 levels=.00] | 0a | . | . | . | . | . | . | 1 | . | . |
| [Limiting_Condition=1.00] | -0.305 | 0.0637 | -0.43 | -0.18 | 22.928 | 1 | 0 | 0.737 | 0.651 | 0.835 |
| [Limiting_Condition=.00] | 0a | . | . | . | . | . | . | 1 | . | . |
| [BMI 2 levels including PNTS and DN=2.00] | -0.166 | 0.0222 | -0.209 | -0.122 | 55.536 | 1 | 0 | 0.847 | 0.811 | 0.885 |
| [BMI 2 levels including PNTS and DN=1.00] | 0a | . | . | . | . | . | . | 1 | . | . |
| [Time=7] * [gender 2 levels including PNS=1.00] | -0.063 | 0.0408 | -0.143 | 0.017 | 2.347 | 1 | 0.126 | 0.939 | 0.867 | 1.018 |
| [Time=7] * [gender 2 levels including PNS=.00] | 0a | . | . | . | . | . | . | 1 | . | . |
| [Time=4] * [gender 2 levels including PNS=1.00] | -0.105 | 0.0365 | -0.177 | -0.034 | 8.343 | 1 | 0.004 | 0.9 | 0.838 | 0.967 |
| [Time=4] * [gender 2 levels including PNS=.00] | 0a | . | . | . | . | . | . | 1 | . | . |
| [Time=2] * [gender 2 levels including PNS=1.00] | -0.028 | 0.0312 | -0.089 | 0.033 | 0.81 | 1 | 0.368 | 0.972 | 0.915 | 1.034 |
| [Time=2] * [gender 2 levels including PNS=.00] | 0a | . | . | . | . | . | . | 1 | . | . |
| [Time=1] * [gender 2 levels including PNS=1.00] | 0a | . | . | . | . | . | . | 1 | . | . |
| [Time=1] * [gender 2 levels including PNS=.00] | 0a | . | . | . | . | . | . | 1 | . | . |

**Supplementary table 21 (Count of PA sessions – Work From Home*Time Interaction Term)**

Tests of Model Effects

| Source | Type III Wald Chi-Square | df | Sig. |
|---|---|---|---|
| (Intercept) | 90.416 | 1 | 0 |
| Time | 34.982 | 3 | 0 |
| gender 2 levels including PNS | 0.415 | 1 | 0.519 |
| Composite_QoL | 48.675 | 1 | 0 |
| age_continuous | 1.837 | 1 | 0.175 |
| Composite occupation and working from home | 4.701 | 2 | 0.095 |
| Binary Living With Others (Alone=1 With Others=0). | 0.048 | 1 | 0.827 |
| Access to a garden Binary Var | 0.224 | 1 | 0.636 |
| Binary Isolation (Total Isolation=1 All Else = 0). | 30.867 | 1 | 0 |
| ethnicity dichotomised including PNS | 0.786 | 1 | 0.375 |
| Composite Housing/Income(0-2) + Education (0-1) Score | 17.488 | 3 | 0.001 |
| Confirmed and suspected Covid-19 2 levels | 0.044 | 1 | 0.835 |
| Limiting_Condition | 23.273 | 1 | 0 |
| BMI 2 levels including PNTS and DN | 55.549 | 1 | 0 |
| Time * Composite occupation and working from home | 9.584 | 6 | 0.143 |

Dependent Variable: PA_Count

| Parameter | B | Std. Error | 95% Wald Confidenc | | Hypothesis Test | | | Exp(B) | 95% Wald Confidenc | |
|---|---|---|---|---|---|---|---|---|---|---|
| | | | Lower | Upper | Wald Chi- | df | Sig. | | Lower | Upper |
| (Intercept) | 1.197 | 0.1434 | 0.916 | 1.478 | 69.706 | 1 | 0 | 3.311 | 2.5 | 4.386 |
| [Time=7] | 0 | 0.0328 | -0.065 | 0.064 | 0 | 1 | 0.995 | 1 | 0.938 | 1.066 |
| [Time=4] | -0.023 | 0.0301 | -0.082 | 0.036 | 0.573 | 1 | 0.449 | 0.977 | 0.922 | 1.037 |
| [Time=2] | 0.051 | 0.0264 | -0.001 | 0.103 | 3.743 | 1 | 0.053 | 1.052 | 0.999 | 1.108 |
| [Time=1] | 0a | . | . | . | . | . | . | 1 | . | . |
| [gender 2 levels including PNS=1.00] | -0.02 | 0.031 | -0.081 | 0.041 | 0.415 | 1 | 0.519 | 0.98 | 0.922 | 1.042 |
| [gender 2 levels including PNS=.00] | 0a | . | . | . | . | . | . | 1 | . | . |
| Composite_QoL | 0.09 | 0.0129 | 0.065 | 0.115 | 48.675 | 1 | 0 | 1.094 | 1.067 | 1.122 |
| age_continuous | 0.002 | 0.0012 | -0.001 | 0.004 | 1.837 | 1 | 0.175 | 1.002 | 0.999 | 1.004 |
| [Composite occupation and working from home=3.00] | -0.02 | 0.0554 | -0.129 | 0.089 | 0.131 | 1 | 0.717 | 0.98 | 0.879 | 1.093 |
| [Composite occupation and working from home=2.00] | 0.021 | 0.0444 | -0.066 | 0.108 | 0.225 | 1 | 0.635 | 1.021 | 0.936 | 1.114 |
| [Composite occupation and working from home=1.00] | 0a | . | . | . | . | . | . | 1 | . | . |
| [Binary Living With Others (Alone=1 With Others=0).=1.00] | -0.009 | 0.0433 | -0.094 | 0.075 | 0.048 | 1 | 0.827 | 0.991 | 0.91 | 1.078 |
| [Binary Living With Others (Alone=1 With Others=0).=.00] | 0a | . | . | . | . | . | . | 1 | . | . |
| [Access to a garden Binary Var=1.00] | 0.016 | 0.0334 | -0.05 | 0.081 | 0.224 | 1 | 0.636 | 1.016 | 0.952 | 1.085 |
| [Access to a garden Binary Var=.00] | 0a | . | . | . | . | . | . | 1 | . | . |
| [Binary Isolation (Total Isolation=1 All Else = 0).=1.00] | -0.303 | 0.0545 | -0.41 | -0.196 | 30.867 | 1 | 0 | 0.739 | 0.664 | 0.822 |
| [Binary Isolation (Total Isolation=1 All Else = 0).=.00] | 0a | . | . | . | . | . | . | 1 | . | . |
| [ethnicity dichotomised including PNS=1.00] | -0.055 | 0.0615 | -0.175 | 0.066 | 0.786 | 1 | 0.375 | 0.947 | 0.839 | 1.068 |
| [ethnicity dichotomised including PNS=.00] | 0a | . | . | . | . | . | . | 1 | . | . |
| [Composite Housing/Income(0-2) + Education (0-1) Score=3.00] | 0.23 | 0.099 | 0.036 | 0.424 | 5.414 | 1 | 0.02 | 1.259 | 1.037 | 1.529 |
| [Composite Housing/Income(0-2) + Education (0-1) Score=2.00] | 0.19 | 0.0984 | -0.002 | 0.383 | 3.749 | 1 | 0.053 | 1.21 | 0.998 | 1.467 |
| [Composite Housing/Income(0-2) + Education (0-1) Score=1.00] | 0.076 | 0.1007 | -0.122 | 0.273 | 0.562 | 1 | 0.453 | 1.078 | 0.885 | 1.314 |
| [Composite Housing/Income(0-2) + Education (0-1) Score=.00] | 0a | . | . | . | . | . | . | 1 | . | . |
| [Confirmed and suspected Covid-19 2 levels=1.00] | -0.005 | 0.0259 | -0.056 | 0.045 | 0.044 | 1 | 0.835 | 0.995 | 0.945 | 1.046 |
| [Confirmed and suspected Covid-19 2 levels=.00] | 0a | . | . | . | . | . | . | 1 | . | . |
| [Limiting_Condition=1.00] | -0.307 | 0.0637 | -0.432 | -0.183 | 23.273 | 1 | 0 | 0.735 | 0.649 | 0.833 |
| [Limiting_Condition=.00] | 0a | . | . | . | . | . | . | 1 | . | . |
| [BMI 2 levels including PNTS and DN=2.00] | -0.166 | 0.0222 | -0.209 | -0.122 | 55.549 | 1 | 0 | 0.847 | 0.811 | 0.885 |
| [BMI 2 levels including PNTS and DN=1.00] | 0a | . | . | . | . | . | . | 1 | . | . |
| [Time=7] * [Composite occupation and working from home=3.00] | -0.127 | 0.0592 | -0.243 | -0.011 | 4.602 | 1 | 0.032 | 0.881 | 0.784 | 0.989 |
| [Time=7] * [Composite occupation and working from home=2.00] | -0.066 | 0.0414 | -0.147 | 0.015 | 2.567 | 1 | 0.109 | 0.936 | 0.863 | 1.015 |
| [Time=7] * [Composite occupation and working from home=1.00] | 0a | . | . | . | . | . | . | 1 | . | . |
| [Time=4] * [Composite occupation and working from home=3.00] | -0.124 | 0.0531 | -0.228 | -0.02 | 5.458 | 1 | 0.019 | 0.883 | 0.796 | 0.98 |
| [Time=4] * [Composite occupation and working from home=2.00] | -0.024 | 0.0371 | -0.096 | 0.049 | 0.405 | 1 | 0.525 | 0.977 | 0.908 | 1.05 |
| [Time=4] * [Composite occupation and working from home=1.00] | 0a | . | . | . | . | . | . | 1 | . | . |
| [Time=2] * [Composite occupation and working from home=3.00] | -0.046 | 0.0456 | -0.135 | 0.044 | 0.996 | 1 | 0.318 | 0.956 | 0.874 | 1.045 |
| [Time=2] * [Composite occupation and working from home=2.00] | -0.038 | 0.0323 | -0.101 | 0.026 | 1.367 | 1 | 0.242 | 0.963 | 0.904 | 1.026 |
| [Time=2] * [Composite occupation and working from home=1.00] | 0a | . | . | . | . | . | . | 1 | . | . |
| [Time=1] * [Composite occupation and working from home=3.00] | 0a | . | . | . | . | . | . | 1 | . | . |
| [Time=1] * [Composite occupation and working from home=2.00] | 0a | . | . | . | . | . | . | 1 | . | . |
| [Time=1] * [Composite occupation and working from home=1.00] | 0a | . | . | . | . | . | . | 1 | . | . |

**Supplementary table 22 (Count of PA sessions – Living with Others*Time Interaction Term)**

Tests of Model Effects

| Source | Type III Wald Chi-Square | df | Sig. |
|---|---|---|---|
| (Intercept) | 91.283 | 1 | 0 |
| Time | 16.548 | 3 | 0.001 |
| gender 2 levels including PNS | 0.433 | 1 | 0.511 |
| Composite_QoL | 49.906 | 1 | 0 |
| age_continuous | 1.738 | 1 | 0.187 |
| Composite occupation and working from home | 3.936 | 2 | 0.14 |
| Binary Living With Others (Alone=1 With Others=0). | 0.002 | 1 | 0.966 |
| Access to a garden Binary Var | 0.206 | 1 | 0.65 |
| Binary Isolation (Total Isolation=1 All Else = 0). | 31.486 | 1 | 0 |
| ethnicity dichotomised including PNS | 0.79 | 1 | 0.374 |
| Composite Housing/Income(0-2) + Education (0-1) Score | 17.628 | 3 | 0.001 |
| Confirmed and suspected Covid-19 2 levels | 0.064 | 1 | 0.8 |
| Limiting_Condition | 22.889 | 1 | 0 |
| BMI 2 levels including PNTS and DN | 54.475 | 1 | 0 |
| Time * Binary Living With Others (Alone=1 With Others=0). | 5.044 | 3 | 0.169 |

| Parameter | B | Std. Error | 95% Wald Confidenc | | Hypothesis Test | | | Exp(B) | 95% Wald Confidenc | |
|---|---|---|---|---|---|---|---|---|---|---|
| | | | Lower | Upper | Wald Chi-! | df | Sig. | | Lower | Upper |
| (Intercept) | 1.232 | 0.1419 | 0.954 | 1.51 | 75.35 | 1 | 0 | 3.428 | 2.595 | 4.527 |
| [Time=7] | -0.064 | 0.0205 | -0.104 | -0.024 | 9.833 | 1 | 0.002 | 0.938 | 0.901 | 0.976 |
| [Time=4] | -0.072 | 0.0181 | -0.107 | -0.036 | 15.752 | 1 | 0 | 0.931 | 0.898 | 0.964 |
| [Time=2] | 0.013 | 0.0155 | -0.017 | 0.043 | 0.725 | 1 | 0.395 | 1.013 | 0.983 | 1.044 |
| [Time=1] | 0a | . | . | . | . | . | . | 1 | . | . |
| [gender 2 levels including PNS=1.00] | -0.02 | 0.031 | -0.081 | 0.04 | 0.433 | 1 | 0.511 | 0.98 | 0.922 | 1.041 |
| [gender 2 levels including PNS=.00] | 0a | . | . | . | . | . | . | 1 | . | . |
| Composite_QoL | 0.091 | 0.0129 | 0.066 | 0.116 | 49.906 | 1 | 0 | 1.095 | 1.068 | 1.123 |
| age_continuous | 0.002 | 0.0012 | -0.001 | 0.004 | 1.738 | 1 | 0.187 | 1.002 | 0.999 | 1.004 |
| [Composite occupation and working from home=3.00] | -0.086 | 0.0488 | -0.182 | 0.009 | 3.125 | 1 | 0.077 | 0.917 | 0.834 | 1.009 |
| [Composite occupation and working from home=2.00] | -0.01 | 0.0404 | -0.09 | 0.069 | 0.066 | 1 | 0.797 | 0.99 | 0.914 | 1.071 |
| [Composite occupation and working from home=1.00] | 0a | . | . | . | . | . | . | 1 | . | . |
| [Binary Living With Others (Alone=1 With Others=0).=1.00] | -0.053 | 0.0503 | -0.151 | 0.046 | 1.102 | 1 | 0.294 | 0.949 | 0.859 | 1.047 |
| [Binary Living With Others (Alone=1 With Others=0).=.00] | 0a | . | . | . | . | . | . | 1 | . | . |
| [Access to a garden Binary Var=1.00] | 0.015 | 0.0334 | -0.05 | 0.081 | 0.206 | 1 | 0.65 | 1.015 | 0.951 | 1.084 |
| [Access to a garden Binary Var=.00] | 0a | . | . | . | . | . | . | 1 | . | . |
| [Binary Isolation (Total Isolation=1 All Else = 0).=1.00] | -0.305 | 0.0544 | -0.412 | -0.199 | 31.486 | 1 | 0 | 0.737 | 0.662 | 0.82 |
| [Binary Isolation (Total Isolation=1 All Else = 0).=.00] | 0a | . | . | . | . | . | . | 1 | . | . |
| [ethnicity dichotomised including PNS=1.00] | -0.055 | 0.0614 | -0.175 | 0.066 | 0.79 | 1 | 0.374 | 0.947 | 0.839 | 1.068 |
| [ethnicity dichotomised including PNS=.00] | 0a | . | . | . | . | . | . | 1 | . | . |
| [Composite Housing/Income(0-2) + Education (0-1) Score=3.00] | 0.23 | 0.0988 | 0.036 | 0.423 | 5.406 | 1 | 0.02 | 1.258 | 1.037 | 1.527 |
| [Composite Housing/Income(0-2) + Education (0-1) Score=2.00] | 0.19 | 0.0982 | -0.003 | 0.382 | 3.73 | 1 | 0.053 | 1.209 | 0.997 | 1.465 |
| [Composite Housing/Income(0-2) + Education (0-1) Score=1.00] | 0.074 | 0.1006 | -0.123 | 0.271 | 0.545 | 1 | 0.461 | 1.077 | 0.884 | 1.312 |
| [Composite Housing/Income(0-2) + Education (0-1) Score=.00] | 0a | . | . | . | . | . | . | 1 | . | . |
| [Confirmed and suspected Covid-19 2 levels=1.00] | -0.007 | 0.0258 | -0.057 | 0.044 | 0.064 | 1 | 0.8 | 0.993 | 0.944 | 1.045 |
| [Confirmed and suspected Covid-19 2 levels=.00] | 0a | . | . | . | . | . | . | 1 | . | . |
| [Limiting_Condition=1.00] | -0.305 | 0.0638 | -0.43 | -0.18 | 22.889 | 1 | 0 | 0.737 | 0.65 | 0.835 |
| [Limiting_Condition=.00] | 0a | . | . | . | . | . | . | 1 | . | . |
| [BMI 2 levels including PNTS and DN=2.00] | -0.164 | 0.0222 | -0.208 | -0.12 | 54.475 | 1 | 0 | 0.849 | 0.813 | 0.887 |
| [BMI 2 levels including PNTS and DN=1.00] | 0a | . | . | . | . | . | . | 1 | . | . |
| [Time=7] * [Binary Living With Others (Alone=1 With Others=0).=1.00] | 0.056 | 0.0503 | -0.043 | 0.154 | 1.234 | 1 | 0.267 | 1.057 | 0.958 | 1.167 |
| [Time=7] * [Binary Living With Others (Alone=1 With Others=0).=.00] | 0a | . | . | . | . | . | . | 1 | . | . |
| [Time=4] * [Binary Living With Others (Alone=1 With Others=0).=1.00] | 0.097 | 0.0459 | 0.008 | 0.187 | 4.51 | 1 | 0.034 | 1.102 | 1.008 | 1.206 |
| [Time=4] * [Binary Living With Others (Alone=1 With Others=0).=.00] | 0a | . | . | . | . | . | . | 1 | . | . |
| [Time=2] * [Binary Living With Others (Alone=1 With Others=0).=1.00] | 0.065 | 0.0395 | -0.012 | 0.143 | 2.735 | 1 | 0.098 | 1.067 | 0.988 | 1.153 |
| [Time=2] * [Binary Living With Others (Alone=1 With Others=0).=.00] | 0a | . | . | . | . | . | . | 1 | . | . |
| [Time=1] * [Binary Living With Others (Alone=1 With Others=0).=1.00] | 0a | . | . | . | . | . | . | 1 | . | . |
| [Time=1] * [Binary Living With Others (Alone=1 With Others=0).=.00] | 0a | . | . | . | . | . | . | 1 | . | . |

#### Measures

##### Outcomes: Physical Activity

**RQ1 & RQ2ii) Muscle-Strengthening Activity (MSA)**

Participant engagement in strength training was measured at each time point with the question, '*In the past month, on average, on how many days per week have you performed strength training?*', with answers ranging from '*0*' to '*4 or more*' *days per week*.

To assess whether participants were meeting WHO recommended levels of muscle strengthening activity, the above score is dichotomised into a binary variable, '*Meeting WHO MSA recommendations'* with 1 representing engaging in 2 or more weekly sessions and 0 representing <2 weekly sessions in line with previous studies with this cohort [15].

**RQ1 & RQ2iii) Moderate/Vigorous Physical Activity (MVPA)**

*Session frequency:* Participant engagement in moderate or vigorous physical activity measured at each time point with the question, '*In the past month, on average, how many times per week have you done 15 minutes or more of moderate or vigorous aerobic physical exercise?*', with answers ranging from '*0'* to '*14 or more'*. Answers of '14 or more' will be treated as 14 times. This is therefore an upper limit constraint set to 2 sessions per day.

*Session duration:* Participants who reported performing at least one session of moderate or vigorous physical activity also reported the average length of their sessions by answering the question, '*In the past month, how long (in minutes) was your average session of moderate or vigorous aerobic physical activity*?'. Answers were recorded on an interval scale which ranges from '*15 minutes'* to '*480 minutes or more'* on a scale increasing in increments of 5 until 60 minutes, and increasing in increments of 10 thereafter. Given the question refers to average session length, the upper limit constraint of 8 hours captures highly atypical and extreme responses.

To estimate whether participants were meeting WHO recommended levels of MVPA training alone, a composite score is derived by multiplying:

*Average session duration x Frequency of Sessions in typical week.*

Possible composite scores are therefore 0 (equivalent to no sessions in a typical week) and thereafter a range of 15 minutes (1 session of the minimum length) to 6720 minutes (14 sessions of 480 minutes).

The above score is dichotomised into a binary variable, '*Meeting WHO MVPA recommendations'* with 1 representing engaging in ≥150 weekly minutes, and 0 representing <150 weekly minutes of moderate/vigorous aerobic physical activity [15].

**RQ1 & RQ2i) Meeting WHO Recommendations**

In addition to identifying those meeting either of the two components of the WHO recommendations respectively, a composite, binary variable, *'Meeting Total WHO Recommended levels of MVPA and MSA'* is produced for each individual at each time point based on the sum of *' Meeting WHO MVPA recommendations'* and *' Meeting WHO MSA recommendations'.* As such a score of 1 corresponds to meeting the requirements for both MSA and MVPA at this time point, and 0 indicates that the participant has failed to meet both recommendations at this time point [15].

<u>**Time-Varying Predictors: Physical and Psychological Wellbeing**</u>

**Quality of Life (Wellbeing, Relationships and Living situation Composite Measure)**
At baseline participants were asked: '*How would you rate the below aspects of your life in the since COVID-19? Living Situation, Social Relationships, Family relationships and Psychological Wellbeing*'. The same question was asked at each follow-up with the question, *'How would you rate the below aspects of your life in the past month?'*. Each of the domains could be scored 1-5, with 1 being poor, 2, 3 average, 4,5 excellent. A continuous mean score would be calculated as:

*Living Situation (1-5) + wellbeing (1-5) + social relationships (1-5) + family relationships (1-5) / (n domains=4)*

**Perceived Risk**
Perceived risk of COVID-19 on a participant's health is reported pre-COVID-19 and at all subsequent time points with the question, '*What risk does COVID-19 pose to your health?'*, with answers ranging from *'Major risk', 'significant risk', 'moderate risk', 'minor risk', 'no risk at all'* and *'don't know'.* dichotomised as *'major to moderate risk*' and '*minimal to no risk and all others'*.

**Overweight**
BMI (derived from baseline reported height and body weight at baseline, 3 and 6 months) dichotomised into overweight or obese (≥25.00) and all others including '*don't know'* and '*prefer not to say*'.

A weighted mean of body weight at baseline and 3 months follow-up will be calculated to estimate BMI at 1 month.

**Confirmed/suspected COVID-19 infection**
Binary yes/no variable derived from the question, *'Do you think you have or had COVID-19?'.* Possible answers included: *'I definitely HAVE COVID-19', 'I think I HAVE COVID-19', 'I definitely HAD COVID-19', 'I think I HAD COVID-19', 'I do not have or think I have had COVID-19', 'Prefer not to say'* and '*Don't know'.* Answers indicating having had or having suspected COVID-19, will be scored as 1. All other responses will be coded as 0.

For one, three and six months follow-up arms only, when testing was more widely available, the above question was preceded by the question: *"Have you been tested for COVID-19?"* with responses: '*Yes and tested positive', 'Yes and tested negative', 'Yes awaiting results' 'No'* and '*Prefer not to say'.* For these three time points therefore, only individual's either awaiting a test or who have never received a test on this variable are then asked the previous question. For homogeneity in this question across the baseline and follow-up arms, those with i) a positive coronavirus test ii) those without a test, but who respond '*I think I have' or 'I think I have had COVID-19'* will be scored as 1. A negative test and all other answers including '*prefer not to say'* and *'don't know'* at these time points will be scored as 0.

<u>**Time-Varying Predictors: Lockdown Situation**</u>

**Employed From Home**
Employment is reported at all time points with the question, '*What is your main occupation at the moment?'.* Being unemployed will be treated as answers of *'Laid off during COVID-19', 'Unemployed*

*since before COVID-19', 'Retired', 'Homemaker', 'Full-time parent or carer', 'Unable to work due to disability' or 'Other'* will be considered unemployed. Answers of '*Employed (full or part-time)', 'Self-employed (full or part-time)', 'Student' and* '*Furloughed during COVID-19)*' will be considered as employed.

Participants who answered as *employed (full or part-time), self-employed (full or part-time), student, furloughed during COVID-19* were also asked, '*Can you do your work or study from home?*', with responses '*Yes, I can do all the work or study from home*', '*No, my work or study cannot be done at home*' and '*I can only do some work or study from home'*.

A composite variable will be derived with 3 levels. Employed from home (Employed AND Yes, I can do all the work or study from home, I can only do some work or study from home) will score 2. Employed not from home (Employed AND No, my work or study cannot be done at home) will be scored 1. Unemployed will be scored 0.

**Isolation Status (Time Variant)**
At all time-points, isolation is measured with the question, *'What is your current isolation status?'*, with possible response options of '*Total isolation/quarantine (not leaving the house for ANY reasons*', '*Some isolation (not leaving the house EXCEPT to buy essential items', 'General isolation but still go out to work*' and *'No isolation (I am free to leave the house whenever I like)'. 'Total isolation/quarantine'* will be scored as a 1 as exercising outdoors even during UK lockdown was permitted for 1 hour. *'Some isolation', 'General isolation but still go out to work'* & *'No isolation'* will be scored as 0.

**<u>Time-Invariant Predictors: Physical and Psychological Wellbeing</u>**

**Pre-COVID-19 Exercise Levels**
Derived in the same way as primary physical activity outcome.
Meeting WHO recommended levels of MSA only before COVID-19 coded as yes/no.

Meeting WHO recommended levels of MVPA only before COVID-19 coded as yes/no.

Meeting WHO recommended levels of MSA and MVPA before COVID-19 coded as yes/no.

**Physical Health Condition Limiting Physical Activity**
At baseline only, participants reported having a physical health condition which might impact their ability to engage in physical activity with the questions: *'Did a doctor or health professional ever tell you that you had any of the following conditions?*' Including: *Heart disease, Stroke, High blood pressure, Hypertension, Diabetes, Dementia, Liver disease, Cancer (within last 5 years), Kidney disease, Lung disease (asthma or COPD), Have undergone organ Transplant, Other conditions leading to immunosuppression (e.g. HIV) or Prefer not to say*.

Participants are also asked: '*Do you now suffer from any condition that limits you from engaging in physical activity, including walking or doing housework?'.* A composite variable of both questions will be summarised in a single binary, 'yes/no' variable with a score of 1 if participants report having any limiting condition or not. This response will be considered fixed for the study period.

**<u>Time-Invariant Predictors: Lockdown Situation</u>**

**Exercise Space Access**
Having access to a garden is a binary 'yes/no' variable asked once at baseline with the question, *'Do you have access to a garden* or *balcony big enough to exercise comfortably?'.* This response will be considered as fixed across the study period.

**Living Alone (Loneliness)**
Binary variable of living alone is measured at baseline with the question, '*How many persons other than yourself (including children) live with you now in the same flat or house*?'. Living alone will be scored as 1 and living with others will be scored as 0.

<u>**Time-Invariant Predictors: Sociodemographic**</u>

**Age**
This will be kept as continuous in years, and will be derived from a free text age field and the drop down age variable, capped at 130.

**Gender**
Binary measure of gender collected at baseline with *Female* coded as 1, all else as 0.

**Ethnicity**
Binary measure of ethnicity collected baseline with *Any White Ethnicity* coded as 1, any other background as 0.

**Composite Socioeconomic Index**
Composite sum score of dummy variables derived from educational level, income and housing tenure with a final range of 0-2.

Education levels are scored as 0 for *No formal qualification, GCSE/School certificate/O-level/CSE, Vocational qualifications (e.g. NVQ1+2) and 1 for A-level/Higher school certificate or equivalent (e.g. NVQ3), Other*, *Bachelor degree or equivalent (e.g.NVQ4)*, M*asters/PhD/PGCE or equivalent* or above.

Housing and Income are a combined measure assessed with the questions, *'What is your housing tenure?'* with possible answers *Owned outright, Mortgage Rented from local authority, Rented from private landlord, Belongs to housing association, Shared ownership (part owned, part rented), Other.*

Income is assessed with the question, *'What was your household annual income BEFORE COVID-19?'* up to 13 499 GBP, 13 500-24 999 GBP, 25 000-49 999 GBP, ≥50 000 GBP, prefer not to say.

A score of 2 is given to Income of 50 000+ GBP AND Housing which is Owned outright or with a mortgage. A score of 1 is given to housing which is Owned outright or with a mortgage AND all other incomes (<50,000+ GBP or Prefer not to say). A score of 0 is for all other incomes AND all other housing. A combined sum score is attained by:
*Composite Socioeconomic Index = Education (0/1) + Income/Housing Tenure (0-2)*.

**Country of Living***
Reported by participants at baseline in response to '*Which country do you live in?'* with options, '*England', 'Wales', 'Scotland', 'Northern Ireland'* or '*Elsewhere'.*
**Included for descriptive purposes only.*
